# Disentangling heterogeneity in Substance Use Disorders: Insights from Genome-Wide Polygenic Scores

**DOI:** 10.1101/2023.11.11.23298413

**Authors:** Laura Vilar-Ribó, Silvia Alemany, Judit Cabana-Domínguez, Natalia Llonga, Lorena Arribas, Lara Grau-López, Constanza Daigre, Bru Cormand, Noèlia Fernàndez-Castillo, Josep Antoni Ramos-Quiroga, María Soler Artigas, Marta Ribasés

**Affiliations:** Psychiatric Genetics Unit, Group of Psychiatry, Mental Health and Addiction, Vall d’Hebron Research Institute (VHIR), Universitat Autònoma de Barcelona, Barcelona, Spain; Department of Mental Health, Hospital Universitari Vall d’Hebron, Barcelona, Spain; Biomedical Network Research Centre on Mental Health (CIBERSAM), Madrid, Spain; Department of Psychiatry and Forensic Medicine, Universitat Autònoma de Barcelona, Barcelona, Spain; Addiction and Dual Diagnosis Unit, Department of Psychiatry, Hospital Universitari Vall d’Hebron, Barcelona, Spain; Department of Genetics, Microbiology, and Statistics, Faculty of Biology, Universitat de Barcelona, Barcelona, Spain; Biomedical Network Research Centre on Rare Diseases (CIBERER), Instituto de Salud Carlos III, Madrid, Spain; Sant Joan de Déu Research Institute (IRSJD), Esplugues de Llobregat, Catalonia, Spain; Institute of Biomedicine of the University of Barcelona (IBUB), Barcelona, Catalonia, Spain

## Abstract

Substance use disorder (SUD) is a global health problem with significant impact on individuals and society. The presentation of SUD is diverse, involving various substances, ages at onset, comorbid conditions, and disease trajectories. Current treatments for SUD struggle to address this heterogeneity, resulting in high relapse rates. SUD often co-occurs with other psychiatric and mental-health related conditions that contribute to the heterogeneity of the disorder and predispose to adverse disease trajectories. Family and genetic studies highlight the role of genetic and environmental factors in the course of SUD, and point to a shared genetic liability between SUDs and comorbid psychopathology. In this study, we aimed to disentangle SUD heterogeneity using a deeply phenotyped SUD cohort and polygenic scores (PGSs) for psychiatric disorders and related traits. We explored associations between PGSs and various SUD-related phenotypes, as well as PGS-environment interactions using information on lifetime emotional, physical and/or sexual abuse. Our results revealed different patterns of associations between the genetic liability for mental-health related traits and SUD-related phenotypes, which may help explain part of the heterogeneity observed in SUD. In our SUD sample, we found associations linking the genetic liability for ADHD with lower educational attainment, the genetic liability for PTSD with higher rates of unemployment, the genetic liability for educational attainment with lower rates of criminal records and unemployment and the genetic liability for well-being with lower rates of outpatient treatments and fewer problems related to family and social relationships. We also found evidence of PGS-environment interactions showing that genetic liability for suicide attempt worsened the psychiatric status in SUD individuals with a history of emotional physical and/or sexual abuse. Collectively, these data contribute to a better understanding of the role of the genetic liability for mental health-related conditions and adverse life experiences in SUD heterogeneity.

## Introduction

Substance use disorder (SUD) is a growing global health problem impacting the individual’s life and the society as a whole. In 2019, 3.2 million people died due to SUD-related causes with 300.000 deaths due to drug or alcohol overdose [1].

The presentation of SUD is highly heterogeneous across a wide range of phenotypic outcomes such as type of substance(s) [2], age at onset of SUD [3,4], individual personality profiles [5,6], presence of comorbid conditions [7] and disease trajectory [8]. For instance, polysubstance use, present in approximately 50% of individuals with SUD [9], has been associated with poorer treatment outcomes [10], higher rates premature death due to overdose [2] and higher rates of mental-health problems and risky behaviours [9]. Early onset substance users are at higher risk for psychosocial problems [3], unemployment [11], low educational attainment [4] and heavier drug abuse in adulthood [8]. The presence of comorbid psychiatric disorders has been associated with adverse disease trajectory, such as poorer treatment adherence in individuals with comorbid major depression disorder or Attention-Deficit Hyperactivity disorder (ADHD)[10,12], increased rates of suicide in individuals with comorbid schizophrenia [13], and worse physical and mental health in individuals with comorbid Post-Traumatic Stress Disorder (PTSD) [14]. In addition, behavioural traits, such as neuroticism, have been associated with lower rates of abstinence and increased symptom severity [6]. Most available inpatient and outpatient treatments for SUD, however, are not well suited to accommodate the observed heterogeneity, resulting in high rates of early treatment termination and relapse [15].

Twin and adoption studies support the role of moderate to high (30-70%) genetic influence on SUD [16] and genome-wide associations studies (GWASs) have identified risk loci associated with substance-specific SUDs [17–20]. These studies, together with other genetic approaches, point to a shared genetic liability and a unitary genetic architecture of SUD across different substances [21,22]. In addition, SUD genetic liability, which can be assessed using polygenic scores (PGSs), presents substantial overlap with psychiatric disorders and behavioural traits [23], and shows the strongest genetic correlations with ADHD, PTSD, anxiety, schizophrenia, depression, bipolar disorder and risk-taking behaviours [22,24,25]. Supporting this idea, a recent study in a deeply phenotyped SUD sample reported that PGSs for substance-specific SUDs were associated with their primary substance-related phenotypes but also with major depression disorder, PTSD, lifetime trauma assessment, being suspended from school or family history of SUD [26]. This finding suggests that the genetic liability for co-occurring psychopathology may explain part of the heterogeneity found in SUD.

In addition, there is growing evidence that the effect of genetic risk on SUD can be moderated by environmental factors, which may also contribute to the individual differences in addictive behaviours [27]. For instance, adverse life experiences, such as trauma exposure or peer drug use, seem to moderate the effect of PGS for cannabis use on lifetime cannabis use [28], the effect of PGS for bipolar disorder on alcohol misuse [29] and the effect of PGS for alcohol problems in adults on earlier alcohol problems [30]

In the present study, we aim to disentangle SUD heterogeneity in a SUD cohort of 1427 individuals who underwent deep phenotyping by conducting a systematic investigation of associations between 39 SUD-related phenotypes and the genetic liability for psychiatric disorders and related traits using PGSs, and to assess whether the profile of PGS associations across SUD-related phenotypes is modulated by exposure to lifetime emotional, physical and/or sexual abuse.

## Materials and methods

### Sample description

A total of 1427 individuals with SUD were recruited at the Addiction and Dual Diagnosis Unit of Hospital Universitari Vall d’Hebron, Barcelona, Spain. Inclusion criteria were age over 18 years old, substance abuse or dependence according to the DSM-IV criteria, European ancestry and a signed informed consent prior to participation. The project was approved by the Ethics Committee at the Hospital Universitari Vall d’Hebron.

### Clinical assessment

The clinical assessment was conducted by trained psychiatrists and psychologists in two different steps: (i) At recruitment, a questionnaire designed ad hoc was administered to gather information on sociodemographic status (sex, age, educational attainment, employment status and criminal record), lifetime medical conditions, psychiatric and SUD family history and substance use related variables (substance(s) of use and/or abuse, age at onset of use, age at onset of SUD, years of substance use and SUD treatment history); (ii) The follow-up interviews were divided into four sessions to evaluate SUD severity, DSM-IV axis I and axis II disorders, heath-related quality of life and personality traits with different scales and questionnaires (Figure 1), detailed below.

**Figure 1.**
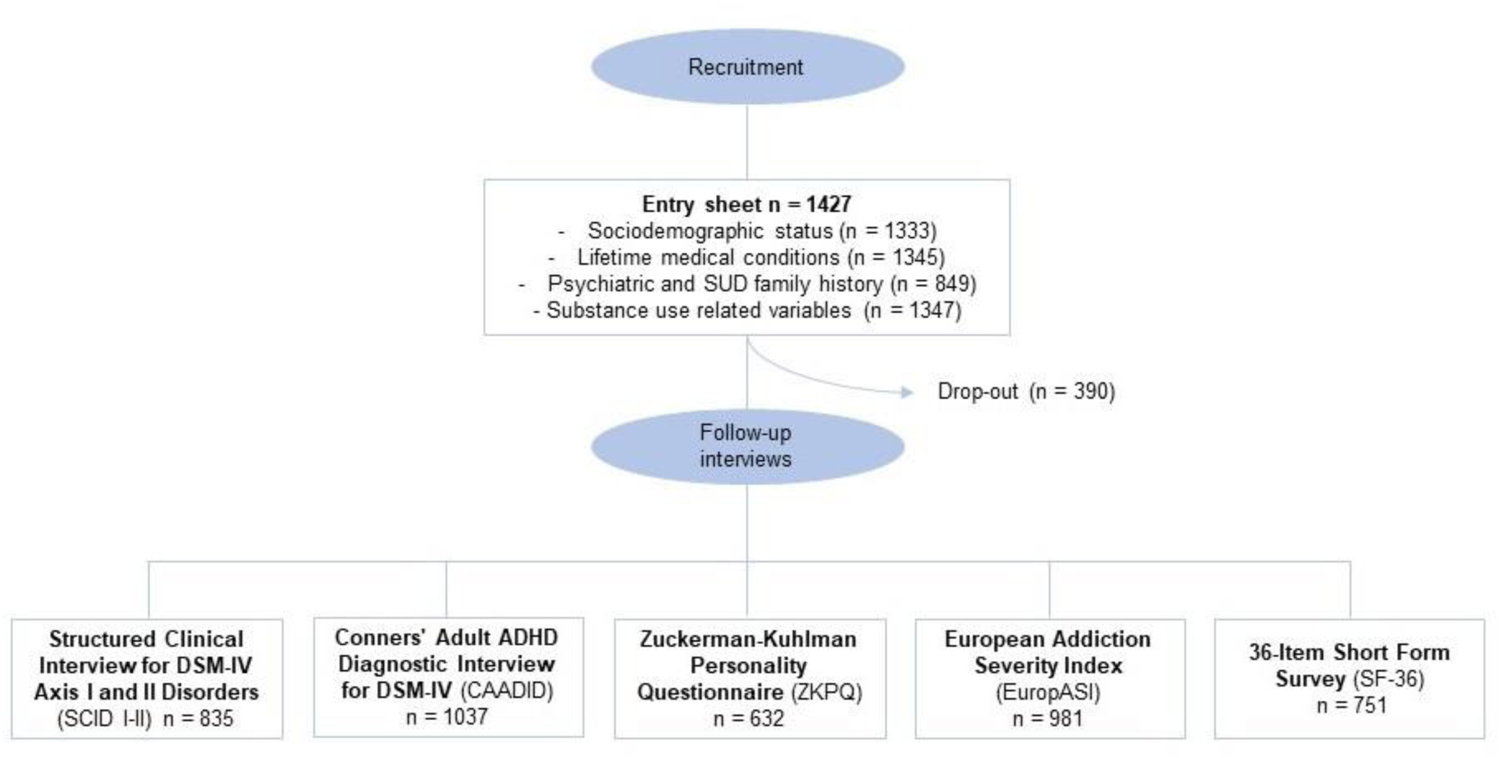
Flowchart. Flowchart with the different stages of the study including the recruitment and follow-up of the SUD sample. Sample size (n) refers to the number of individuals with at least one item of the interview available.

The structured Clinical Interview for Axis I and II Disorders of the DSM-IV (SCID-I and SCID-II) [31] and the Conners’ Adult ADHD diagnostic interview for DSM-IV (CAADDID-II) [32] were used to assess psychiatric comorbidity. The Spanish version of the Zuckerman–Kuhlman Personality Questionnaire (ZKPQ) [33] was used to assess personality features including neuroticism-anxiety, activity, sociability, impulsive sensation-seeking and aggression-hostility. The validated Spanish version of the European Addiction Severity Index interview (EuropASI) [34] is design to provide information about aspects of an individual’s life which may contribute to his/her substance abuse, specifically on the following areas: legal status, employment status, medical status, psychiatric status, drug use, alcohol use and family/social relationships. Scores ranging from 0 to 1 are estimated, with higher scores indicating greater severity. The 36-Item Short Form Survey (SF-36) was administered to measure the self-reported health and quality of life, both physically and mentally with higher scores indicating better health [35]. After data curation, 39 phenotypes with sample size >300 were considered and classified into three categories: SUD variables (n= 8), comorbidity and personality traits (n=15) and sociodemographic and health outcomes (n=16) (Table 1). For binary traits, with n1 individuals in one group and n2 individuals in the other group, effective sample size was calculated with the formula 4/(1/n1+1/n2).

**Table 1.** Summary of the 39 SUD-related phenotypes.

| **Phenotypes** | ***n*<sup>a</sup>** | **Summary** |
| --- | --- | --- |
| Sex: Male (%) | 1092 | 76.5 |
| Age; Mean (SD) | 1427 | 38.6 (10.3) |
| **SUD variables** |  |  |
| Age at onset of substance use; Mean (SD) | 1347 | 17.05 (1.36) |
| Age at onset of SUD; Mean (SD) | 1339 | 19.32 (1.4) |
| Years between substance use and SUD; Median (IQR) | 1325 | 1 (3) |
| Years of substance use as proportion of lifespan; Median (IQR) | 1148 | 34 (33) |
| Number of substances consumed | 869 |  |
| 1 (%) |  | 23.2 |
| 2 (%) |  | 29.5 |
| 3 or more (%) |  | 47.3 |
| Number of therapeutic community interventions; Median (IQR) | 1314 |  |
| 0 (%) |  | 64.5 |
| 1 (%) |  | 22.7 |
| 2 (%) |  | 7.1 |
| 3 or more (%) |  | 5.7 |
| Number of inpatient detoxifications; Median (IQR) | 1327 |  |
| 0 (%) |  | 70.5 |
| 1 (%) |  | 16.0 |
| 2 (%) |  | 6.8 |
| 3 or more (%) |  | 6.7 |
| Number of outpatient treatments; Median (IQR) | 1252 |  |
| 0 (%) |  | 28.5 |
| 1 (%) |  | 37.9 |
| 2 (%) |  | 16.6 |
| 3 or more (%) |  | 17 |
| **Comorbidity and personality traits** |  |  |
| *Mental disorders in DSM-IV* |  |  |
| Borderline personality disorder (%) | 447 | 15.9 |
| Major depressive disorder (%) | 828 | 37.8 |
| Antisocial personality disorder (%) | 560 | 21.3 |
| Psychotic disorder (%) | 593 | 7.8 |
| Anxiety disorder (%) | 654 | 24.9 |
| Attention deficit hyperactivity disorder (%) | 700 | 21.5 |
| *Zuckerman–Kuhlman Personality Questionnaire (ZKPQ)* |  |  |
| Neuroticism Anxiety personality factor; Mean (SD) | 663 | 10.8 (4.9) |
| Aggression Hostility personality factor; Mean (SD) | 667 | 8.93 (3.15) |
| Sociability personality factor; Mean (SD) | 632 | 6.6 (3.4) |
| Impulsive sensation seeking personality factor; Mean (SD) | 666 | 10.5 (4.3) |
| Activity personality factor; Mean (SD) | 665 | 8.09 (3.5) |
| Suicide attempt (%) | 618 | 47.8 |
| Suicide ideation (%) | 731 | 30.1 |
| Psychotic symptoms (%) | 1281 | 58.6 |
| Sleeping disturbances (%) | 1228 | 54.6 |
| **Sociodemographic and health outcomes** |  |  |
| *EuropASI* |  |  |
| Legal status; Median (IQR) | 984 | 0 (.1) |
| Employment status; Median (IQR) | 984 | .55 (.5) |
| Medical status; Median (IQR) | 982 | 0 (.1) |
| Psychiatric status; Median (IQR) | 984 | .4 (.3) |
| Drug use; Median (IQR) | 984 | .2 (.2) |
| Alcohol use; Median (IQR) | 984 | .1 (.3) |
| Family/Social relationships; Median (IQR) | 981 | .4 (.5) |
| *36-Item Short Form Survey (SF-36)* |  |  |
| Physical health; Mean (SD) | 751 | 48.4 (1.8) |
| Mental health; Mean (SD) | 751 | 35.5 (13.7) |
| Criminal record (%) | 713 | 43.1 |
| Unemployment (%) | 1057 | 73.4 |
| Number of psychiatric hospitalizations; Median (IQR) | 760 |  |
| 0 (%) |  | 79.9 |
| 1 (%) |  | 9.9 |
| 2 (%) |  | 4.5 |
| 3 or more (%) |  | 5.7 |
| Psychiatric family history (%) | 840 | 44.8 |
| Lifetime medical conditions (%) | 1334 | 54.4 |
| Substance use family history (%) | 818 | 59.6 |
| Educational attainment | 1333 |  |
| 1 (Incomplete primary school) (%) |  | 14.5 |
| 2 (Primary school) (%) |  | 40.9 |
| 3 (Secondary/High school) (%) |  | 35.6 |
| 4 (Bachelor's degree or higher) (%) |  | 9.1 |

<sup>a</sup> For binary traits, with n1 individuals in one group and n2 individuals in the other group, effective sample size was calculated with the formula $4/(1/n1+1/n2)$ .

### Genotyping and quality control

Genomic DNA was isolated from whole blood by the salting-out procedure and genotyped with the Illumina Infinium Global Screening Array-24 version 2 (GSA v2) (Illumina, CA, San Diego, USA) in two different waves (434 and 993 samples, respectively). Pre-imputation quality control was done with the PLINK 2.0 software [36] and included individual and variant filtering based on the following parameters: variant call rate >0.95 (before individual filtering), individual call rate >0.98, autosomal heterozygosity deviation (| Fhet | <0.2), variant call rate >0.98 (after individual filtering), difference in variant missingness between cases and controls <0.02, SNP Hardy-Weinberg equilibrium (HWE) (*p* >1e−06 in controls or *p* >1e−10 in cases) and minor allele frequency (MAF) > 0.01. Genetic outliers were identified by principal component analysis (PCA) using PLINK 2.0 and the mixed ancestry 1000G reference panel [37]. Non-European individuals were excluded if their principal component (PC) values for PC1 and PC2 were greater than 1 standard deviation from the mean-centring point for the study population. Related and duplicated samples were identified by the “KING-robust kinship estimator” analysis in PLINK 2.0 [38] and one individual was excluded from each pair of subjects with kinship coefficient > 0.0442. Imputation was done with McCarthy tools, for data preparation, and the Michigan Imputation Server [39], using the Haplotype Reference Consortium (HRC Version r1.1 2016) reference panel (GRCh37/hg19). Variants were excluded in case of allele mismatch between the reference panel and the study dataset (chi2>900). Post-imputation dosage files with imputation INFO score >0.8 and MAF >0.01 were considered for subsequent analyses.

### Polygenic scores

Polygenic scores (PGSs) were constructed in our using the PRS-CS software [40], PLINK 2.0 and available GWASs summary statistics (**Table S1**), on psychiatric disorders (ADHD [41] anxiety (http://www.nealelab.is/uk-biobank/), bipolar disorder [42], depression [43], post-traumatic Stress Disorder (PTSD) [44] and schizophrenia [45]), behavioural traits (risk tolerance [46], suicide attempt [47]) and other related traits (educational attainment [48] and well-being [49]). PGSs were computed and standardized to a mean of 0 and a standard deviation of 1 for all disorders and traits.

### Statistical analysis

#### Association between polygenic scores and SUD-related phenotypes

The profile of PGSs associations across the SUD-related phenotypes were assessed with the appropriate regression models depending on the nature of the outcome variable with R: logistic regression for binary variables, linear regression for continuous variables, ordinal regression for ordinal categorical variables and negative binominal for count variables. Prior to the analysis, logarithmic transformations were applied to continuous variables not following a normal distribution (“age at onset of substance use” and “age at onset of SUD”). Additionally, linear regression residuals were checked for continuous variables with significant results to ensure they followed a normal distribution. Age, sex, genotyping batch and the 10 first PCs were included as covariates in all analyses. Additionally, for the variables “age at onset of substance use”, “age at onset of SUD”, “years between substance use and SUD”, and “years of substance use as a proportion of lifespan”, the main drug of use, abuse or dependence was included as a covariate. *P*-values were corrected for multiple comparisons using PhenoSpD [50,51], a command line R based tool for estimating phenotypic correlations and multiple testing correction. The effective number of independent variables estimated was 35 using the VeffLi model and the corrected *p*-value threshold was set at *p*-value < 1.46e-03 [52].

#### Interaction between polygenic scores and emotional, physical and/or sexual abuse in SUD-related phenotypes

For those PGSs associated with any outcome, interaction with emotional, physical and/or sexual abuse was tested in a subset of 735 individuals who had completed the EuropASI family/social relationships questionnaire and information on emotional, physical and/or sexual abuse was available. Potential interaction effects were tested introducing an interaction term (PGS*abuse) in the regression model adjusted for age, sex, genotyping batch and the 10 first PCs as covariates. Multiple comparison corrected *p-*value, calculated with PhenoSpD in R, was set at *p* < 2.05e-03 [50,51]. For significant interactions, PGS-outcome associations were stratified by exposure to emotional, physical and/or sexual abuse.

## Results

Our cohort consisted of 1427 individuals (76.5% male), with a mean age of 38.6 years (SD = 10.3) (**Table 1**). The vast majority of subjects were polysubstance users and 47% fulfilled SUD criteria for three or more substances.

### Polygenic scores for psychiatric disorders

After multiple testing correction we found significant associations between the PGS for ADHD and lower educational attainment (OR=0.85, 95% CI [0.93, 0.77], *p*=1.20e-03) and between the PGS for PTSD and unemployment (OR=1.23, 95% CI [1.09, 1.4], *p*=1.00e-03) **(Figure 2, Table S2a and S2e)**.

**Figure 2.**
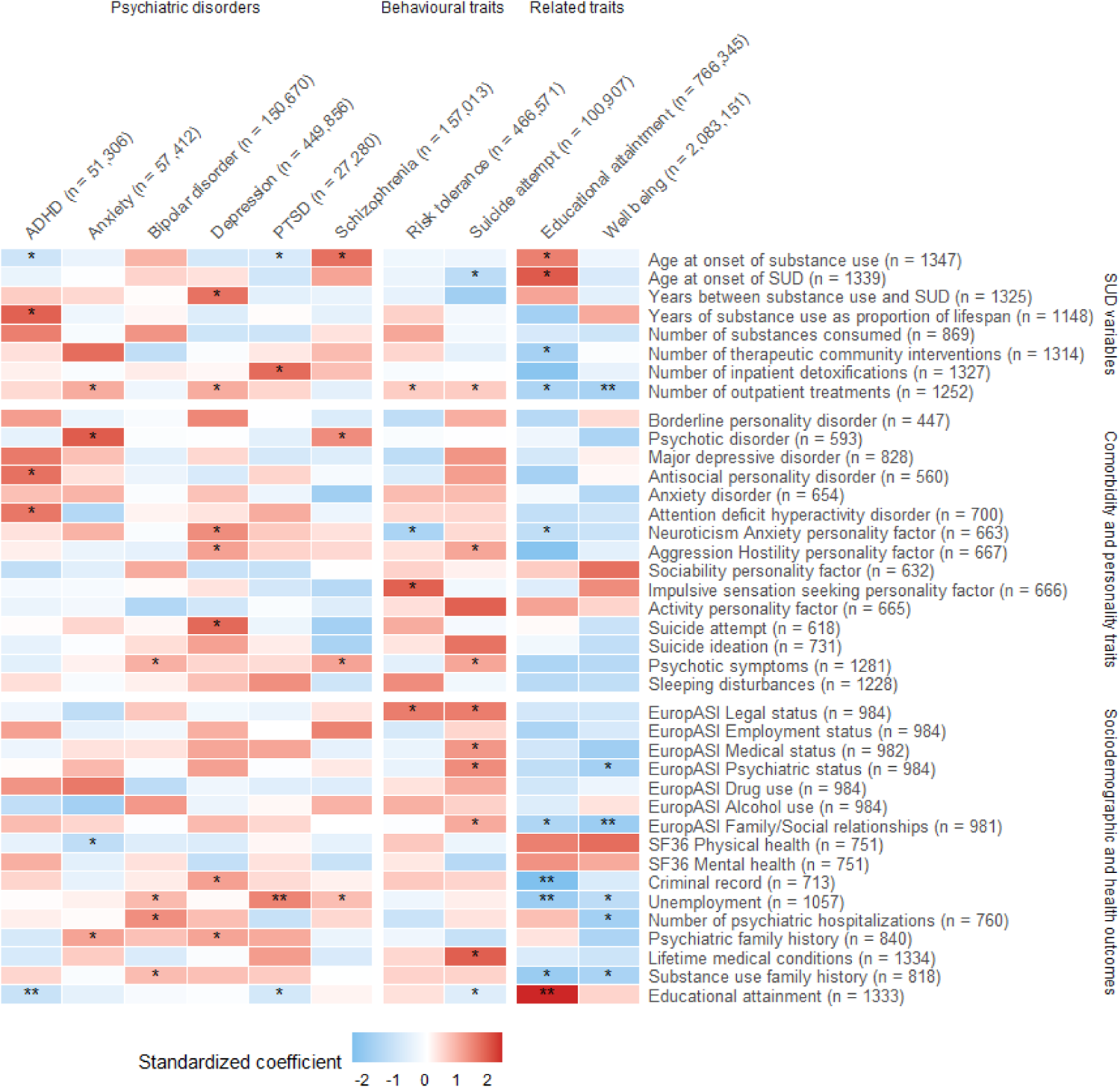
Heatmap for the results of the association between PGSs and SUD-related phenotypes. Association pattern between 10 PGSs for psychiatric disorders, behavioural and related traits with the SUD-related phenotypes; *nominal significance *p-*values; \*\**p-*values that passed multiple testing correction using PhenoSpD (P<1.46e-03). ADHD = Attention-deficit hyperactivity disorder; PTSD = Post-traumatic Stress Disorder. Standardized coefficient corresponds to Beta for continuous variables, log(OR) for binary and ordinal variables and log(IRR) for count variables.

Despite not surpassing the multiple testing correction threshold (*p* < 0.05), we found additional nominal associations. PGS for ADHD was associated with early-onset of first substance use (Beta (β)=-0.01, 95% CI [-0.03, -3.00e-04]), longer term substance use as a proportion of the lifespan (β=1.07, 95% CI [0.03, 2.11]), lifetime diagnosis of ADHD (OR=1.24, 95% CI [1.06, 1.45]), and antisocial personality disorder (OR=1.24, 95% CI [1.04, 1.47]) **(Figure 2, Table S2a)**. PGS for anxiety was associated with more outpatient treatments (incidence rate ratio (IRR)=1.09, 95% CI [1.03, 1.15]), psychotic disorders across the lifetime (OR=1.49, 95% CI [1.16, 1.92]), poorer self-perceived physical health status measured with the SF-36 instrument (β=-0.93, 95% CI [-1.64, -0.22]) and psychiatric family history (OR=1.16, 95% CI [1.01, 1.33]) **(Figure 2, Table S2b)**. PGS for bipolar disorder was associated with higher rates of psychotic symptoms (OR=1.14, 95% CI [1.02, 1.28]), unemployment (OR=1.14, 95% CI [1.01, 1.29]), psychiatric hospitalizations (IRR=1.26, 95% CI [1.03, 1.53]) and substance use family history (OR=1.15, 95% CI [1.0, 1.32]) **(Figure 2, Table S2c)**. PGS for depression showed association with slower transition from substance use to SUD (IRR=1.1, 95% CI [1.01, 1.21]), more outpatient treatments (IRR=1.09, 95% CI [1.03, 1.15]) and higher rates of neuroticism-anxiety (β=0.36, 95% CI [4e-3, 0.72]) and aggression-hostility (β=0.27, 95% CI [0.04, 0.5]) according to the ZKPQ, suicide attempts (OR=1.18, 95% CI [1.02, 1.37]), criminal records (OR=1.27, 95% CI [1.09, 1.48]) and psychiatric family history (OR=1.16, 95% CI [1.01, 1.32]) **(Figure 2, Table S2d)**. PGS for PTSD was associated with early-onset of first substance use (β=-0.01, 95% CI [-0.03, -1e-3]), more inpatient detoxifications (IRR=1.13, 95% CI [1.01, 1.27]), and lower educational attainment (OR=0.87, 95% CI [0.97, 0.79]) **(Figure 2, Table S2e)**. And lastly, PGS for schizophrenia was associated with later onset of substance use (β=0.01, 95% CI [3e-4, 0.03]), psychotic disorders (OR=1.37, 95% CI [1.05, 1.78]), higher rates of psychotic symptoms (OR=1.15, 95% CI [1.03, 1.29]) and unemployment (OR=1.13, 95% CI [1, 1.28]) **(Figure 2, Table S2f)**.

### Polygenic scores for behavioural traits

None of the associations between GPSs for behavioural traits and SUD-related phenotypes surpassed multiple testing correction, however, nominally significant associations (*p* < 0.05) are detailed bellow.

PGS for risk tolerance showed associations with more outpatient treatments (IRR=1.06, 95% CI [1, 1.12]), lower rates of neuroticism-anxiety (β=-0.51, 95% CI [-0.88, – 0.14]) and higher rates of impulsive sensation seeking (β=0.32, 95% CI [3e-3, 0.65]) according to the ZKPQ and higher rates of legal problems measured by the EuropASI index (OR=1.18, 95% CI [1.02, 1.36]) **(Figure 2, Table S2g)**. PGS for suicide attempt was associated with early-onset of SUD (β=-0.02, 95% CI [-0.04, -0.01]), more outpatient treatments (IRR=1.06, 95% CI [1, 1.12]), higher rates of aggression-hostility (β=0.26, 95% CI [0.02, 0.51]) and psychotic symptoms (OR=1.15, 95% CI [1.03, 1.29]), more legal (OR=1.18, 95% CI [1.01, 1.37]), medical (OR=1.14, 95% CI [1, 1.29]), psychiatric (OR=1.13, 95% CI [1.01, 1.27]) and family/social (OR=1.13, 95% CI [1, 1.26]) problems measured by the EuropASI index, more lifetime medical conditions (OR=1.14, 95% CI [1, 1.29]) and lower educational attainment (OR=0.90, 95% CI [1, 0.81]) **(Figure 2, Table S2h)**.

### Polygenic scores for educational attainment and well-being

After multiple testing correction we found significant associations between PGS for educational attainment and less criminal records (OR=0.67, 95% CI [0.57, 0.78], *p*=8.03e-07), unemployment (OR=0.82, 95% CI [0.71, 0.92], *p*=1.34e-03) and higher educational attainment (OR=1.39, 95% CI [1.54, 1.25], *p*=3.34e-10), as well as associations between PGS for well-being and less outpatient treatments (OR=0.91, 95% CI [0.86, 0.96], *p*=7.00e-04) and family/social problems (OR=0.83, 95% CI [0.74, 0.93], *p*= 1.00e-03) **(Figure 2, Table S2i and S2j)**.

Nominal associations (*p* < 0.05) include the association between PGS for educational attainment and later onset of substance use (β=0.02, 95% CI [0.01, 0.03]) and SUD (β=0.02, 95% CI [4e-3, 0.04]), less therapeutic community interventions (OR=0.89, 95% CI [0.81, 0.98]) or outpatient treatments (OR=0.91, 95% CI [0.87, 0.97]), lower rates of neuroticism-anxiety (β=-0.4, 95% CI [-0.78, -0.01] 2), less social-familiar problems (OR=0.86, 95% CI [0.77, 0.96]), and substance use family history (OR=0.80, 95% CI [0.7, 0.92]) **(Figure 2, Table S2i)**. Moreover, PGS for well-being showed association with lower rates of psychiatric problems (OR=0.87, 95% CI [0.78, 0.97]), less unemployment (OR=0.88, 95% CI [0.77, 0.99]), psychiatric hospitalizations (OR=0.78, 95% CI [0.68, 0.95]), and substance use family history (OR=0.83, 95% CI [0.77, 0.99]) **(Figure 2, Table S2j)**.

### Interaction between polygenic scores and emotional, physical and/or sexual abuse on SUD-related phenotypes

Information on lifetime emotional, physical and/or sexual abuse was available for a total of 735 individuals with SUD, 45.6% of which (n=335) reported having experienced some sort of abuse across their lifetime. PGS*abuse interaction analysis was performed for those PGSs nominally associated with any outcome **(Table S3).** We found one significant interaction where lifetime abuse moderates the association between PGS for suicide attempt and the psychiatric status measured by the EuropASI index (OR=1.35, 95% CI [1.03, 1.78], *p*=2.94e-02). Specifically, the genetic liability for suicide attempt was associated with worse psychiatric status scores among those having experienced lifetime emotional, physical and/or sexual abuse (OR=1.33 95% CI [0.48, 0.09], *p*= 4.67e-04), while the association was not significant for those not exposed (**Figure 3**).

**Figure 3.**
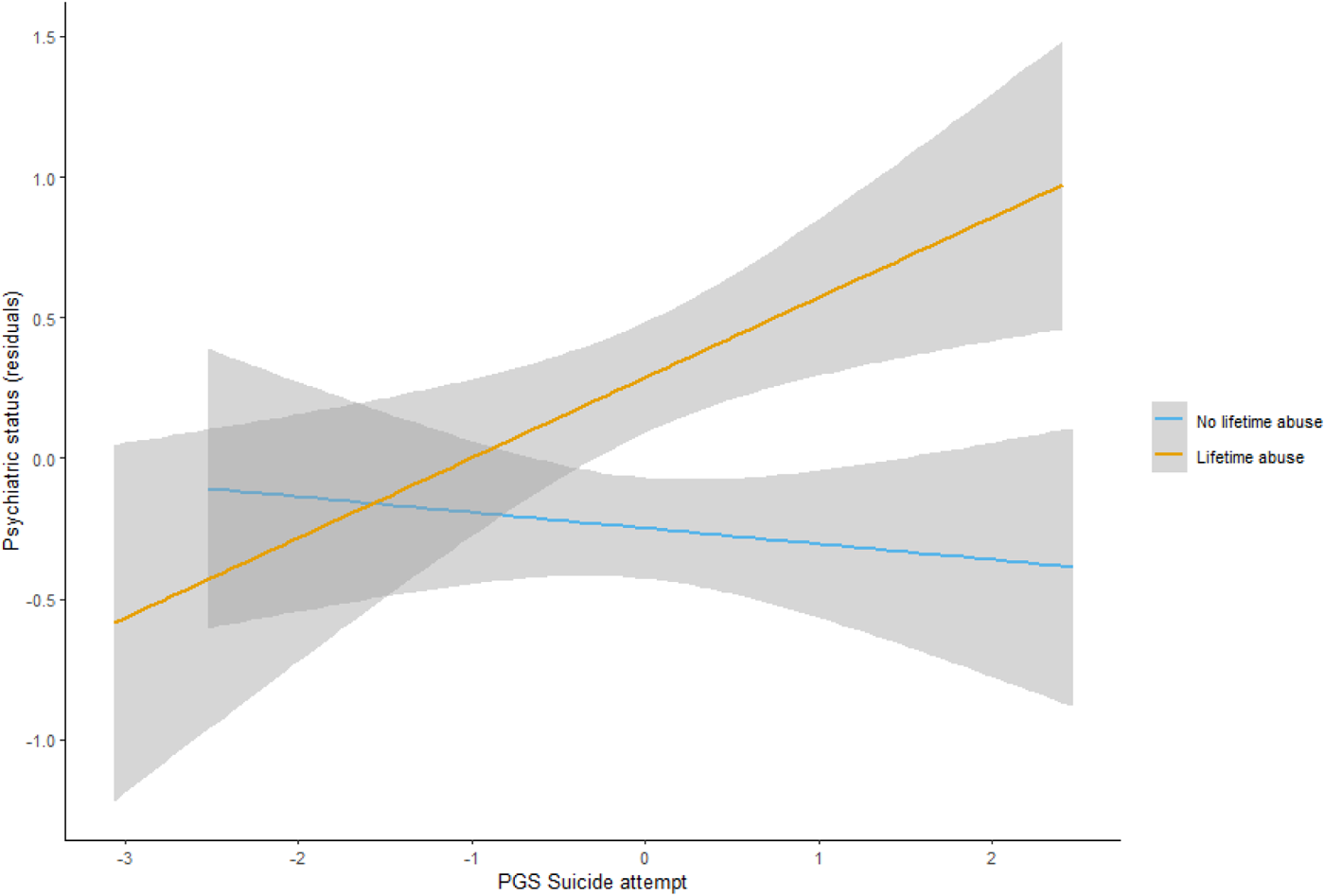
Statistically significant result from the interaction analysis. Interaction between PGS for suicide attempt and lifetime emotional, physical and/or sexual abuse in psychiatric status measured by the EuropASI index. The X axis presents the PGS for suicide attempt, and the Y axis shows the residuals from an ordinal regression model with psychiatric status as outcome adjusted for age, sex, genotyping batch and the 10 first principal components for those individuals who suffered lifetime abuse (in yellow) and those who did not (in blue).

## Discussion

There is immense clinical and genetic heterogeneity among individuals with SUD, and current treatment approaches fail to accommodate this variability, resulting in poor treatment adherence and high rates of relapse [53]. In this study, we utilized multidimensional data from a deeply phenotyped SUD cohort and individual genetic liability information for a broad range of mental-health related traits using PGSs, to provide new insights into the heterogeneity of the disorder. Our approach included the systematic association of 10 PGSs for psychiatric disorders, behavioural and other related traits with 39 SUD-related phenotypes, and the assessment of PGS-environmental interactions using information on emotional, physical and/or sexual abuse. Our main findings suggest that the genetic liability for ADHD, PTSD and suicide attempt, in conjunction with environmental factors, may underlie, at least partially, the observed heterogeneity in SUD-related phenotypes such as educational attainment, unemployment and psychiatric status.

PGSs analysis on the SUD-related phenotypes builds on previous findings supporting links between the genetic risk for psychiatric disorders and a wide variety of SUD outcomes. In line with this evidence, our results suggest that the genetic liability for mental-health related traits exhibits different patterns of associations with SUD-related phenotypes. Specifically, we replicated previous findings linking the genetic liability for ADHD with lower educational attainment [54], the genetic liability for PTSD with higher rates of unemployment [55], and the genetic liability for higher educational attainment with lower rates of criminal records and unemployment [56,57]. While these associations were described in the general population, our results suggest that these patterns remain in individuals with SUD. Moreover, our findings showed that the genetic liability for well-being is associated with better outcomes, namely lower rates of outpatient treatments and fewer problems related to family and social relationships, which is consistent with the role of the genetics underlying well-being in healthy family relationships [58].

Despite not surpassing multiple comparison correction, we found evidence supporting previously reported associations. For instance, PGSs for ADHD, schizophrenia and educational attainment were associated with their respective primary phenotype, confirming the validity of the approach. In addition, we identified an association between the genetic liability for depression and higher rates of suicide attempts. This is consistent with previous findings linking PGSs for depression with suicide attempt [59–62], and studies suggesting an increased risk of suicide attempt and ideation among individuals with comorbid SUD and major depressive disorder [63,64]. Our findings add to the evidence supporting that genetic liability for depression may have a relevant role regarding suicide attempt and ideation in the context of SUD.

Our results also shed light into the association between the genetic liability for multiple psychiatric disorders and poor SUD-related outcomes. These include early age at onset of substance use and high number of outpatient treatments, strengthening the notion that genetic susceptibility to psychiatric diseases and behavioural traits may play a role in promoting the initiation and impeding the cessation of substance use [12,65–67]. For instance, we found that individuals with higher PGS for ADHD showed earlier onset and more prolonged substance use, while those with higher PGS for depression showed faster transition from substance use to SUD and more outpatient treatments. Similarly, individuals with higher PGS for PTSD showed earlier onset of substance use and more inpatient treatments.

Moreover, PGSs for educational attainment or suicide attempt were associated with multiple outcomes (more than ten). Increased genetic risk for educational attainment was associated with less therapeutic interventions, late age at onset of substance use or SUD and less SUD family history or problems related with family and social relationships. These findings are consistent with previous evidence showing that the genetic liability for education attainment is linked to decreased SUD severity [68] and a recent study by Kinreich et al., (2021) suggesting that polygenic liability to years of education could be used to predict remission in patients with alcohol use disorder. Additionally, the genetic liability for suicide attempt showed the strongest association with early age at onset of SUD, number of outpatient treatments, higher rates of psychotic symptoms, and a wide range of medical, psychological and legal problems. Adding to this evidence, we report a significant interaction between PGS for suicide attempt and having been exposed to lifetime emotional, physical or sexual abuse in the psychiatric status of SUD individuals. While it is well established that exposure to sexual trauma and/or abuse increases the risk for substance use and mental health problems later in life [70], we found that the genetic liability for suicide attempt exacerbates the negative impact on mental health problems in individuals with a history of abuse. Similar findings have been reported for cannabis use [28] or bipolar disorder [71], where exposure to trauma and/or maltreatment potentiates the polygenic risk for these disorders. Overall, these results highlight that focusing on exposed individuals may render genetic effects that may not be found when environmental exposures are not considered.

Although many results are well supported by prior research, we also found that for some disorders PGSs were not associated with their primary phenotype. For instance, PGSs for anxiety or depression did not show an association with anxiety disorder or major depressive disorder in the SUD dataset. The reasons for this lack of association may include selection bias, complex relationships between SUD and comorbid conditions and limited sample size for some of the outcomes. Moreover, PGS for suicide attempt was not associated with suicide behaviours, namely suicide ideation and attempt, in our SUD dataset. Suicide attempt is a clinically complex phenotype that can vary greatly in frequency and intensity [72]. Even though the GWAS meta-analysis used to construct PGS for suicide attempt aimed to harmonize data across various cohorts by including clinical samples from major psychiatric disorders and individuals from the Million Veterans Project sample [47], differences in population characteristics or assessment methods of the phenotype may account for the inconclusive results observed in our dataset. In previous studies, reliability of PGS-based predictions of suicide attempt has been inconsistent when applied to independent datasets [73–75], and Lannoy et al., (2022) found evidence for the interaction between PGS for suicide attempt and drug use on suicide ideation. Together, these results highlight the multifactorial nature of suicide attempt and suggest that other factors, such as psychiatric comorbidity, SUD type or severity and environmental factors, should be taken into account when assessing suicide risk.

It is important to be cautious when comparing results from PGSs for the disorders and traits tested, taking into consideration the variations in statistical power between some of them. The differences in sample size among the GWAS meta-analyses used to construct PGSs, as well as among the outcomes, could have contributed to the uneven pattern of associations observed. Additionally, other factors such as environmental factors and sex differences may play a significant role in certain aspects of SUD heterogeneity. Furthermore, our results suggest that the patterns of lifetime comorbidity in SUD result, in part, from the contribution of genetic factors. However, it is currently unknown whether substance use is a consequence of underlying psychiatric disorders or whether it increases the risk of mental health problems later in life. Access to longitudinal data would provide new and valuable information to assess causal relationships between SUD and comorbid conditions and to examine the impact of the genetic liability on disease progression.

This study supports that the genetic liability for distinct mental-health related traits plays a role in the heterogeneity of SUD and can influence disease outcome in terms of severity, comorbidity rates and socio-demographic factors. There is also evidence for PGS-environment interactions between the genetic liability for suicide attempt and lifetime emotional physical and/or sexual abuse on the psychiatric status of individuals with SUD. These results encourage the use of PGSs and gene-environment interactions to better understand the heterogeneity of SUD and complex traits.

## Author Contributions

L.V.R., and L.A. participated in the DNA isolation and preparation of samples. L.V.R., M.S.A. and M.R. undertook the statistical analyses. L.V.R., S.A., J.C.D., N.L., M.S.A., and M.R. contributed to the interpretation of data for the work. L.G.L., and C.D., contributed to the clinical assessment and recruitment of patients. J.A.R.Q. participated in the study design, clinical assessment and coordination of the clinical research. M.R. and M.S.A. conceived the project, wrote the protocol and coordinated the study design and the statistical analyses. M.S.A. and M.R. supervised the project and the manuscript preparation. All authors revised the work critically for important intellectual content and have approved the final version.

## Data Availability

Materials and analysis code for this study are available by emailing upon request

