## Supplementary tables 1, 2 and 3 for "Disentangling heterogeneity in Substance Use Disorders: Insights from Genome-Wide Polygenic Scores"

**Supplementary Table 1.**

*Discovery GWASs of psychiatric diseases, behavioral and related traits used to construct the GPSs*

| Trait | n cases | n controls | n total [a] | *ref* |
|---|---|---|---|---|
| Attention-Deficit Hyperactivity Disorder | 20,183 | 35,191 | 51,306 | [1] |
| Anxiety | 16,730 | 101,021 | 57,412 | http://www.nealelab.is/uk-biobank/ |
| Bipolar Disorder | 41,917 | 371,549 | 150,670 | [2] |
| Depression | 170,756 | 329,443 | 449,856 | [3] |
| Post-Traumatic Stress Disorder | 9,354 | 25,175 | 27,280 | [4] |
| Schizophrenia | 67390 | 94015 | 157,013 | [5] |
| Risk tolerance | - | - | 466,571 | [6] |
| Suicide attempt | 26,590 | 492,022 | 100,907 | [7] |
| Educational attainment | - | - | 766,345 | [8] |
| Well being | - | - | 2,083,151 | [9] |

[a] For binary traits effective sample size was calculated with the formula 4/(1/n cases+1/n controls)

**Supplementary Table 2a.** Association between the GPS for ADHD and 39 clinical variables from the SUD phenome. In bold nominal significant results.

| Phenotypes | *n* [a] | Regression | Estimate [b] | 95% CI | *p* |
|---|---|---|---|---|---|
| **SUD-related phenotypes** | | | | | |
| Age at onset of substance use [c] | 1308 | Linear | -0,01 | -0,03 , -3,00E-04 | **4,53E-02** |
| Age at onset of SUD [c] | 1280 | Linear | -0,01 | -0,02 , 0,01 | 0,39 |
| Years between substance use and SUD | 1280 | Linear | 1,04 | 0,95 , 1,14 | 0,36 |
| Years of substance use as proportion of lifespan | 1199 | Linear | 1,07 | 0,03 , 2,1 | **4,42E-02** |
| Number of substances consumed | 879 | Negative binomial | 1,03 | 0,99 , 1,07 | 0,19 |
| Number of therapeutic community interventions | 1345 | Negative binomial | 1,01 | 0,92 , 1,11 | 0,80 |
| Number of inpatient detoxifications | 1327 | Negative binomial | 1,03 | 0,92 , 1,15 | 0,62 |
| Number of outpatient treatments | 1252 | Negative binomial | 1,05 | 0,99 , 1,11 | 0,09 |
| **Comorbidity and personality traits** | | | | | |
| *Mental disorders in DSM-IV* | | | | | |
| Borderline personality disorder | 447 | Logistic | 1,12 | 0,92 , 1,36 | 0,28 |
| Major depressive disorder | 828 | Logistic | 1,10 | 0,96 , 1,27 | 0,17 |
| Antisocial personality disorder | 560 | Logistic | 1,24 | 1,04 , 1,47 | **1,60E-02** |
| Psychotic disorder | 593 | Logistic | 1,02 | 0,79 , 1,3 | 0,90 |
| Anxiety disorder | 654 | Logistic | 1,03 | 0,88 , 1,2 | 0,71 |
| Attention deficit hyperactivity disorder | 700 | Logistic | 1,24 | 1,06 , 1,45 | **7,35E-03** |
| *Zuckerman–Kuhlman Personality Questionnaire (ZKPQ)* | | | | | |
| Neuroticism Anxiety personality factor | 663 | Linear | 0,07 | -0,29 , 0,43 | 0,72 |
| Aggression Hostility personality factor | 667 | Linear | 0,15 | -0,08 , 0,38 | 0,21 |
| Sociability personality factor | 632 | Linear | -0,18 | -0,44 , 0,08 | 0,18 |
| Impulsive sensation seeking personality factor | 666 | Linear | 0,01 | -0,3 , 0,32 | 0,94 |
| Activity personality factor | 665 | Linear | 0,02 | -0,24 , 0,28 | 0,88 |
| Suicide attempt | 618 | Logistic | 1,03 | 0,89 , 1,19 | 0,67 |
| Suicide ideation | 731 | Logistic | 0,98 | 0,84 , 1,15 | 0,80 |
| Psychotic symptoms | 1281 | Logistic | 1,00 | 0,89 , 1,12 | 0,98 |
| Sleeping disturbances | 1228 | Logistic | 1,02 | 0,91 , 1,14 | 0,76 |

| | | | | | |
|---|---|---|---|---|---|
| **Sociodemographic and health phenotypes** | | | | | |
| *EuropASI* | | | | | |
| Legal status | 984 | Ordinal | 1,00 | 0,87 , 1,16 | 0,97 |
| Employment status | 984 | Ordinal | 1,08 | 0,97 , 1,2 | 0,18 |
| Medical status | 982 | Ordinal | 1,03 | 0,92 , 1,17 | 0,60 |
| Psychiatric status | 984 | Ordinal | 1,01 | 0,91 , 1,13 | 0,86 |
| Drug use | 984 | Ordinal | 1,06 | 0,95 , 1,19 | 0,28 |
| Alcohol use | 984 | Ordinal | 0,96 | 0,86 , 1,08 | 0,54 |
| Family/Social relationships | 981 | Ordinal | 1,10 | 0,99 , 1,23 | 0,09 |
| *36-Item Short Form Survey (SF-36)* | | | | | |
| Physical health | 751 | Linear | -0,52 | -1,23 , 0,19 | 0,15 |
| Mental health | 751 | Linear | 0,49 | -0,45 , 1,44 | 0,31 |
| Criminal record | 713 | Logistic | 1,13 | 0,98 , 1,32 | 0,10 |
| Unemployment | 1057 | Logistic | 1,03 | 0,91 , 1,17 | 0,61 |
| Number of psychiatric hospitalizations | 760 | Negative binomial | 1,05 | 0,86 , 1,29 | 0,61 |
| Psychiatric family history | 840 | Logistic | 0,97 | 0,85 , 1,11 | 0,66 |
| Lifetime medical conditions | 1334 | Logistic | 0,99 | 0,88 , 1,11 | 0,84 |
| Substance use family history | 818 | Logistic | 1,10 | 0,96 , 1,27 | 0,16 |
| Educational attainment | 1333 | Ordinal | 0,85 | 0,93 , 0,77 | **1,20E-03** |

[a] For binary traits sample size was calculated with the formula 4/(1/n1+1/n2)

[b] OR is reported for logistic regression and ordinal regression; Beta is reported for lineal regression;  IRR is reported for negative binomial regression

[c] Logarithmic transformations were applied to continuous variables not following a normal distribution

**Supplementary Table 2b**. Association between the GPS for anxiety and 39 clinical variables from the SUD phenome. In bold nominal significant results.

| Phenotypes | *n* [a] | Regression | Estimate [b] | 95% CI | *p* |
|---|---|---|---|---|---|
| **SUD-related phenotypes** | | | | | |
| Age at onset of substance use [c] | 1308 | Linear | -0,01 | -0,02 , 0,01 | 0,36 |
| Age at onset of SUD [c] | 1280 | Linear | 0,00 | -0,02 , 0,01 | 0,77 |
| Years between substance use and SUD | 1280 | Linear | 1,04 | 0,95 , 1,13 | 0,45 |
| Years of substance use as proportion of lifespan | 1199 | Linear | -0,03 | -1,07 , 1,02 | 0,96 |
| Number of substances consumed | 879 | Negative binomial | 1,01 | 0,97 , 1,05 | 0,79 |
| Number of therapeutic community interventions | 1345 | Negative binomial | 1,10 | 1 , 1,21 | 0,05 |
| Number of inpatient detoxifications | 1327 | Negative binomial | 1,01 | 0,9 , 1,13 | 0,84 |
| Number of outpatient treatments | 1252 | Negative binomial | 1,09 | 1,03 , 1,15 | **2,67E-03** |
| **Comorbidity and personality traits** | | | | | |
| *Mental disorders in DSM-IV* | | | | | |
| Borderline personality disorder | 447 | Logistic | 1,02 | 0,84 , 1,24 | 0,86 |
| Major depressive disorder | 828 | Logistic | 1,03 | 0,9 , 1,19 | 0,63 |
| Antisocial personality disorder | 560 | Logistic | 1,09 | 0,92 , 1,29 | 0,30 |
| Psychotic disorder | 593 | Logistic | 1,49 | 1,16 , 1,92 | **1,93E-03** |
| Anxiety disorder | 654 | Logistic | 1,04 | 0,89 , 1,21 | 0,63 |
| Attention deficit hyperactivity disorder | 700 | Logistic | 0,86 | 0,74 , 1,01 | 0,06 |
| *Zuckerman–Kuhlman Personality Questionnaire (ZKPQ)* | | | | | |
| Neuroticism Anxiety personality factor | 663 | Linear | 0,23 | -0,12 , 0,59 | 0,20 |
| Aggression Hostility personality factor | 667 | Linear | 0,07 | -0,16 , 0,3 | 0,56 |
| Sociability personality factor | 632 | Linear | -0,08 | -0,34 , 0,17 | 0,53 |
| Impulsive sensation seeking personality factor | 666 | Linear | 0,02 | -0,29 , 0,33 | 0,91 |
| Activity personality factor | 665 | Linear | 0,04 | -0,22 , 0,29 | 0,79 |
| Suicide attempt | 618 | Logistic | 1,07 | 0,93 , 1,23 | 0,37 |
| Suicide ideation | 731 | Logistic | 1,01 | 0,87 , 1,18 | 0,88 |
| Psychotic symptoms | 1281 | Logistic | 1,06 | 0,95 , 1,19 | 0,28 |
| Sleeping disturbances | 1228 | Logistic | 0,98 | 0,88 , 1,1 | 0,77 |

| | | | | | |
|---|---|---|---|---|---|
| **Sociodemographic and health phenotypes** | | | | | |
| *EuropASI* | | | | | |
| Legal status | 984 | Ordinal | 0,92 | 0,8 , 1,07 | 0,28 |
| Employment status | 984 | Ordinal | 0,97 | 0,87 , 1,08 | 0,57 |
| Medical status | 982 | Ordinal | 1,08 | 0,96 , 1,22 | 0,23 |
| Psychiatric status | 984 | Ordinal | 1,08 | 0,97 , 1,2 | 0,16 |
| Drug use | 984 | Ordinal | 1,08 | 0,97 , 1,21 | 0,17 |
| Alcohol use | 984 | Ordinal | 0,95 | 0,85 , 1,06 | 0,36 |
| Family/Social relationships | 981 | Ordinal | 1,07 | 0,96 , 1,19 | 0,26 |
| *36-Item Short Form Survey (SF-36)* | | | | | |
| Physical health | 751 | Linear | -0,93 | -1,64 , -0,22 | **1,07E-02** |
| Mental health | 751 | Linear | 0,06 | -0,89 , 1,01 | 0,90 |
| Criminal record | 713 | Logistic | 0,94 | 0,81 , 1,1 | 0,47 |
| Unemployment | 1057 | Logistic | 1,05 | 0,93 , 1,19 | 0,44 |
| Number of psychiatric hospitalizations | 760 | Negative binomial | 1,03 | 0,84 , 1,27 | 0,77 |
| Psychiatric family history | 840 | Logistic | 1,16 | 1,01 , 1,33 | **4,16E-02** |
| Lifetime medical conditions | 1334 | Logistic | 1,06 | 0,95 , 1,19 | 0,31 |
| Substance use family history | 818 | Logistic | 1,02 | 0,88 , 1,17 | 0,80 |
| Educational attainment | 1333 | Ordinal | 0,92 | 1,02 , 0,83 | 0,10 |

[a] For binary traits sample size was calculated with the formula 4/(1/n1+1/n2)

[b] OR is reported for logistic regression and ordinal regression; Beta is reported for lineal regression; IRR is reported for negative binomial regression

[c] Logarithmic transformations were applied to continuous variables not following a normal distribution

**Supplementary Table 2c**. Association between the GPS for bipolar disorder and 39 clinical variables from the SUD phenome. In bold nominal significant results.

| Phenotypes | *n* [a] | Regression | Estimate [b] | 95% CI | *p* |
|---|---|---|---|---|---|
| **SUD-related phenotypes** | | | | | |
| Age at onset of substance use [c] | 1308 | Linear | 0,01 | -0,01 , 0,02 | 0,33 |
| Age at onset of SUD [c] | 1280 | Linear | 0,00 | -0,02 , 0,02 | 0,90 |
| Years between substance use and SUD | 1280 | Linear | 1,01 | 0,93 , 1,11 | 0,78 |
| Years of substance use as proportion of lifespan | 1199 | Linear | 0,20 | -0,85 , 1,24 | 0,71 |
| Number of substances consumed | 879 | Negative binomial | 1,02 | 0,98 , 1,07 | 0,26 |
| Number of therapeutic community interventions | 1345 | Negative binomial | 0,92 | 0,84 , 1,01 | 0,09 |
| Number of inpatient detoxifications | 1327 | Negative binomial | 1,03 | 0,92 , 1,16 | 0,59 |
| Number of outpatient treatments | 1252 | Negative binomial | 0,99 | 0,94 , 1,05 | 0,85 |
| **Comorbidity and personality traits** | | | | | |
| *Mental disorders in DSM-IV* | | | | | |
| Borderline personality disorder | 447 | Logistic | 1,03 | 0,85 , 1,26 | 0,74 |
| Major depressive disorder | 828 | Logistic | 0,94 | 0,81 , 1,08 | 0,37 |
| Antisocial personality disorder | 560 | Logistic | 1,02 | 0,86 , 1,21 | 0,85 |
| Psychotic disorder | 593 | Logistic | 1,08 | 0,84 , 1,4 | 0,53 |
| Anxiety disorder | 654 | Logistic | 0,98 | 0,83 , 1,15 | 0,78 |
| Attention deficit hyperactivity disorder | 700 | Logistic | 1,04 | 0,89 , 1,22 | 0,59 |
| *Zuckerman–Kuhlman Personality Questionnaire (ZKPQ)* | | | | | |
| Neuroticism Anxiety personality factor | 663 | Linear | -0,07 | -0,45 , 0,3 | 0,71 |
| Aggression Hostility personality factor | 667 | Linear | 0,05 | -0,18 , 0,29 | 0,65 |
| Sociability personality factor | 632 | Linear | 0,16 | -0,11 , 0,43 | 0,24 |
| Impulsive sensation seeking personality factor | 666 | Linear | 0,05 | -0,28 , 0,38 | 0,76 |
| Activity personality factor | 665 | Linear | -0,09 | -0,36 , 0,18 | 0,53 |
| Suicide attempt | 618 | Logistic | 1,04 | 0,9 , 1,2 | 0,62 |
| Suicide ideation | 731 | Logistic | 1,05 | 0,9 , 1,23 | 0,54 |
| Psychotic symptoms | 1281 | Logistic | 1,14 | 1,02 , 1,28 | **2,58E-02** |
| Sleeping disturbances | 1228 | Logistic | 1,00 | 0,9 , 1,12 | 0,95 |

| | | | | | |
|---|---|---|---|---|---|
| **Sociodemographic and health phenotypes** | | | | | |
| *EuropASI* | | | | | |
| Legal status | 984 | Ordinal | 1,09 | 0,95 , 1,26 | 0,22 |
| Employment status | 984 | Ordinal | 0,98 | 0,88 , 1,09 | 0,73 |
| Medical status | 982 | Ordinal | 1,08 | 0,95 , 1,22 | 0,23 |
| Psychiatric status | 984 | Ordinal | 1,00 | 0,9 , 1,11 | 0,94 |
| Drug use | 984 | Ordinal | 0,93 | 0,83 , 1,04 | 0,19 |
| Alcohol use | 984 | Ordinal | 1,04 | 0,93 , 1,16 | 0,48 |
| Family/Social relationships | 981 | Ordinal | 1,01 | 0,9 , 1,12 | 0,92 |
| *36-Item Short Form Survey (SF-36)* | | | | | |
| Physical health | 751 | Linear | -0,62 | -1,36 , 0,12 | 0,10 |
| Mental health | 751 | Linear | 0,32 | -0,67 , 1,31 | 0,53 |
| Criminal record | 713 | Logistic | 1,08 | 0,93 , 1,25 | 0,32 |
| Unemployment | 1057 | Logistic | 1,14 | 1,01 , 1,29 | **3,72E-02** |
| Number of psychiatric hospitalizations | 760 | Negative binomial | 1,26 | 1,03 , 1,53 | **2,23E-02** |
| Psychiatric family history | 840 | Logistic | 1,12 | 0,98 , 1,28 | 0,11 |
| Lifetime medical conditions | 1334 | Logistic | 0,99 | 0,89 , 1,11 | 0,87 |
| Substance use family history | 818 | Logistic | 1,15 | 1 , 1,32 | **4,89E-02** |
| Educational attainment | 1333 | Ordinal | 0,97 | 1,06 , 0,88 | 0,51 |

[a] For binary traits sample size was calculated with the formula 4/(1/n1+1/n2)

[b] OR is reported for logistic regression and ordinal regression; Beta is reported for lineal regression; IRR is reported for negative binomial regression

[c] Logarithmic transformations were applied to continuous variables not following a normal distribution

**Supplementary Table 2d**. Association between the GPS for depression and 39 clinical variables from the SUD phenome. In bold nominal significant results.

| Phenotypes | *n* [a] | Regression | Estimate [b] | 95% CI | *p* |
|---|---|---|---|---|---|
| **SUD-related phenotypes** | | | | | |
| Age at onset of substance use [c] | 1308 | Linear | 0,01 | -0,03 , 2,00E-03 | 0,09 |
| Age at onset of SUD [c] | 1280 | Linear | 0,00 | -0,02 , 0,01 | 0,54 |
| Years between substance use and SUD | 1280 | Linear | 1,01 | 1,01 , 1,21 | **3,49E-02** |
| Years of substance use as proportion of lifespan | 1199 | Linear | 0,20 | -1,24 , 0,86 | 0,72 |
| Number of substances consumed | 879 | Negative binomial | 1,02 | 0,95 , 1,04 | 0,80 |
| Number of therapeutic community interventions | 1345 | Negative binomial | 0,92 | 0,9 , 1,08 | 0,76 |
| Number of inpatient detoxifications | 1327 | Negative binomial | 1,03 | 0,91 , 1,15 | 0,69 |
| Number of outpatient treatments | 1252 | Negative binomial | 0,99 | 1,03 , 1,15 | **3,04E-03** |
| **Comorbidity and personality traits** | | | | | |
| *Mental disorders in DSM-IV* | | | | | |
| Borderline personality disorder | 447 | Logistic | 1,03 | 0,93 , 1,38 | 0,22 |
| Major depressive disorder | 828 | Logistic | 0,94 | 0,88 , 1,17 | 0,87 |
| Antisocial personality disorder | 560 | Logistic | 1,02 | 0,83 , 1,17 | 0,82 |
| Psychotic disorder | 593 | Logistic | 1,08 | 0,85 , 1,41 | 0,50 |
| Anxiety disorder | 654 | Logistic | 0,98 | 0,88 , 1,2 | 0,73 |
| Attention deficit hyperactivity disorder | 700 | Logistic | 1,04 | 0,92 , 1,25 | 0,41 |
| *Zuckerman–Kuhlman Personality Questionnaire (ZKPQ)* | | | | | |
| Neuroticism Anxiety personality factor | 663 | Linear | -0,07 | 4,00E-03 , 0,72 | **4,91E-02** |
| Aggression Hostility personality factor | 667 | Linear | 0,05 | 0,04 , 0,5 | **2,18E-02** |
| Sociability personality factor | 632 | Linear | 0,16 | -0,41 , 0,11 | 0,25 |
| Impulsive sensation seeking personality factor | 666 | Linear | 0,05 | -0,22 , 0,41 | 0,56 |
| Activity personality factor | 665 | Linear | -0,09 | -0,29 , 0,23 | 0,83 |
| Suicide attempt | 618 | Logistic | 1,04 | 1,02 , 1,37 | **3,04E-02** |
| Suicide ideation | 731 | Logistic | 1,05 | 0,95 , 1,3 | 0,20 |
| Psychotic symptoms | 1281 | Logistic | 1,14 | 0,98 , 1,23 | 0,12 |
| Sleeping disturbances | 1228 | Logistic | 1,00 | 0,93 , 1,17 | 0,48 |

| | | | | | |
|---|---|---|---|---|---|
| **Sociodemographic and health phenotypes** | | | | | |
| *EuropASI* | | | | | |
| Legal status | 984 | Ordinal | 1,09 | 0,87 , 1,16 | 0,92 |
| Employment status | 984 | Ordinal | 0,98 | 0,96 , 1,19 | 0,25 |
| Medical status | 982 | Ordinal | 1,08 | 0,99 , 1,27 | 0,06 |
| Psychiatric status | 984 | Ordinal | 1,00 | 1 , 1,23 | 0,06 |
| Drug use | 984 | Ordinal | 0,93 | 0,87 , 1,09 | 0,68 |
| Alcohol use | 984 | Ordinal | 1,04 | 0,88 , 1,11 | 0,86 |
| Family/Social relationships | 981 | Ordinal | 1,01 | 0,99 , 1,23 | 0,08 |
| *36-Item Short Form Survey (SF-36)* | | | | | |
| Physical health | 751 | Linear | -0,62 | -1,35 , 0,09 | 0,09 |
| Mental health | 751 | Linear | 0,32 | -1,09 , 0,84 | 0,80 |
| Criminal record | 713 | Logistic | 1,08 | 1,09 , 1,48 | **1,82E-03** |
| Unemployment | 1057 | Logistic | 1,14 | 0,92 , 1,17 | 0,58 |
| Number of psychiatric hospitalizations | 760 | Negative binomial | 1,26 | 0,95 , 1,4 | 0,15 |
| Psychiatric family history | 840 | Logistic | 1,12 | 1,01 , 1,32 | **3,70E-02** |
| Lifetime medical conditions | 1334 | Logistic | 0,99 | 0,91 , 1,15 | 0,70 |
| Substance use family history | 818 | Logistic | 1,15 | 0,99 , 1,31 | 0,06 |
| Educational attainment | 1333 | Ordinal | 0,97 | 1,09 , 0,88 | 0,69 |

[a] For binary traits sample size was calculated with the formula 4/(1/n1+1/n2)

[b] OR is reported for logistic regression and ordinal regression; Beta is reported for lineal regression;  IRR is reported for negative binomial regression

[c] Logarithmic transformations were applied to continuous variables not following a normal distribution

**Supplementary Table 2e.** Association between the GPS for post-traumatic stress disorder and 39 clinical variables from the SUD phenome. In bold nominal significant results.

| Phenotypes | *n* [a] | Regression | Estimate [b] | 95% CI | *p* |
|---|---|---|---|---|---|
| **SUD-related phenotypes** | | | | | |
| Age at onset of substance use [c] | 1308 | Linear | -0,01 | -0,03 , -1,00E-03 | **3,72E-02** |
| Age at onset of SUD [c] | 1280 | Linear | -0,01 | -0,03 , 0,01 | 0,23 |
| Years between substance use and SUD | 1280 | Linear | 0,98 | 0,9 , 1,08 | 0,70 |
| Years of substance use as proportion of lifespan | 1199 | Linear | 0,16 | -0,88 , 1,21 | 0,76 |
| Number of substances consumed | 879 | Negative binomial | 0,99 | 0,96 , 1,04 | 0,79 |
| Number of therapeutic community interventions | 1345 | Negative binomial | 1,01 | 0,92 , 1,11 | 0,89 |
| Number of inpatient detoxifications | 1327 | Negative binomial | 1,13 | 1,01 , 1,27 | **4,15E-02** |
| Number of outpatient treatments | 1252 | Negative binomial | 1,05 | 1 , 1,11 | 0,08 |
| **Comorbidity and personality traits** | | | | | |
| *Mental disorders in DSM-IV* | | | | | |
| Borderline personality disorder | 447 | Logistic | 1,04 | 0,85 , 1,27 | 0,69 |
| Major depressive disorder | 828 | Logistic | 0,92 | 0,79 , 1,06 | 0,23 |
| Antisocial personality disorder | 560 | Logistic | 1,11 | 0,93 , 1,31 | 0,24 |
| Psychotic disorder | 593 | Logistic | 1,00 | 0,78 , 1,29 | 0,97 |
| Anxiety disorder | 654 | Logistic | 0,96 | 0,82 , 1,13 | 0,63 |
| Attention deficit hyperactivity disorder | 700 | Logistic | 1,15 | 0,99 , 1,35 | 0,07 |
| *Zuckerman–Kuhlman Personality Questionnaire (ZKPQ)* | | | | | |
| Neuroticism Anxiety personality factor | 663 | Linear | 0,15 | -0,22 , 0,52 | 0,43 |
| Aggression Hostility personality factor | 667 | Linear | 0,19 | -0,05 , 0,43 | 0,12 |
| Sociability personality factor | 632 | Linear | -0,16 | -0,43 , 0,11 | 0,24 |
| Impulsive sensation seeking personality factor | 666 | Linear | -0,09 | -0,41 , 0,23 | 0,59 |
| Activity personality factor | 665 | Linear | 0,05 | -0,22 , 0,32 | 0,73 |
| Suicide attempt | 731 | Logistic | 1,04 | 0,88 , 1,22 | 0,68 |
| Suicide ideation | 618 | Logistic | 1,00 | 0,86 , 1,16 | 0,99 |
| Psychotic symptoms | 1281 | Logistic | 1,09 | 0,97 , 1,22 | 0,15 |
| Sleeping disturbances | 1228 | Logistic | 1,08 | 0,97 , 1,22 | 0,17 |

| | | | | | |
|---|---|---|---|---|---|
| **Sociodemographic and health phenotypes** | | | | | |
| *EuropASI* | | | | | |
| Legal status | 984 | Ordinal | 1,00 | 0,86 , 1,15 | 0,96 |
| Employment status | 984 | Ordinal | 1,00 | 0,9 , 1,12 | 0,99 |
| Medical status | 982 | Ordinal | 1,13 | 0,99 , 1,28 | 0,07 |
| Psychiatric status | 984 | Ordinal | 1,00 | 0,9 , 1,12 | 0,99 |
| Drug use | 984 | Ordinal | 0,96 | 0,86 , 1,08 | 0,50 |
| Alcohol use | 984 | Ordinal | 1,00 | 0,9 , 1,13 | 0,94 |
| Family/Social relationships | 981 | Ordinal | 1,06 | 0,95 , 1,19 | 0,28 |
| *36-Item Short Form Survey (SF-36)* | | | | | |
| Physical health | 751 | Linear | 0,32 | -0,65 , 1,29 | 0,52 |
| Mental health | 751 | Linear | -0,21 | -0,94 , 0,52 | 0,57 |
| Criminal record | 713 | Logistic | 1,12 | 0,96 , 1,3 | 0,15 |
| Unemployment | 1057 | Logistic | 1,23 | 1,09 , 1,4 | **1,00E-03** |
| Number of psychiatric hospitalizations | 760 | Negative binomial | 0,86 | 0,7 , 1,06 | 0,15 |
| Psychiatric family history | 840 | Logistic | 1,15 | 1 , 1,32 | 0,05 |
| Lifetime medical conditions | 1334 | Logistic | 1,09 | 0,97 , 1,22 | 0,13 |
| Substance use family history | 818 | Logistic | 1,12 | 0,97 , 1,29 | 0,11 |
| Educational attainment | 1333 | Ordinal | 0,87 | 0,97 , 0,79 | **9,48E-03** |

[a] For binary traits sample size was calculated with the formula 4/(1/n1+1/n2)

[b] OR is reported for logistic regression and ordinal regression; Beta is reported for lineal regression;  IRR is reported for negative binomial regression

[c] Logarithmic transformations were applied to continuous variables not following a normal distribution

**Supplementary Table 2f**. Association between the GPS for schizophrenia and 39 clinical variables from the SUD phenome. In bold nominal significant results.

| Phenotypes | *n* [a] | Regression | Estimate [b] | 95% CI | *p* |
|---|---|---|---|---|---|
| **SUD-related phenotypes** | | | | | |
| Age at onset of substance use [c] | 1308 | Linear | 0,01 | 3,00E-04 , 0,03 | **4,61E-02** |
| Age at onset of SUD [c] | 1280 | Linear | 0,01 | -0,01 , 0,02 | 0,44 |
| Years between substance use and SUD | 1280 | Linear | 0,99 | 0,9 , 1,08 | 0,80 |
| Years of substance use as proportion of lifespan | 1199 | Linear | -0,11 | -1,15 , 0,94 | 0,84 |
| Number of substances consumed | 879 | Negative binomial | 1,01 | 0,97 , 1,06 | 0,58 |
| Number of therapeutic community interventions | 1345 | Negative binomial | 1,04 | 0,95 , 1,14 | 0,43 |
| Number of inpatient detoxifications | 1327 | Negative binomial | 1,06 | 0,95 , 1,19 | 0,28 |
| Number of outpatient treatments | 1252 | Negative binomial | 0,98 | 0,93 , 1,04 | 0,49 |
| **Comorbidity and personality traits** | | | | | |
| *Mental disorders in DSM-IV* | | | | | |
| Borderline personality disorder | 447 | Logistic | 1,00 | 0,82 , 1,22 | 0,99 |
| Major depressive disorder | 828 | Logistic | 0,93 | 0,8 , 1,07 | 0,31 |
| Antisocial personality disorder | 560 | Logistic | 1,04 | 0,87 , 1,23 | 0,67 |
| Psychotic disorder | 593 | Logistic | 1,37 | 1,05 , 1,78 | **1,97E-02** |
| Anxiety disorder | 654 | Logistic | 0,88 | 0,75 , 1,03 | 0,12 |
| Attention deficit hyperactivity disorder | 700 | Logistic | 0,98 | 0,84 , 1,15 | 0,84 |
| *Zuckerman–Kuhlman Personality Questionnaire (ZKPQ)* | | | | | |
| Neuroticism Anxiety personality factor | 663 | Linear | 0,07 | -0,3 , 0,44 | 0,71 |
| Aggression Hostility personality factor | 667 | Linear | 0,18 | -0,06 , 0,42 | 0,14 |
| Sociability personality factor | 632 | Linear | 0,01 | -0,26 , 0,28 | 0,94 |
| Impulsive sensation seeking personality factor | 666 | Linear | -0,17 | -0,49 , 0,16 | 0,31 |
| Activity personality factor | 665 | Linear | -0,01 | -0,27 , 0,26 | 0,97 |
| Suicide attempt | 618 | Logistic | 0,90 | 0,78 , 1,04 | 0,17 |
| Suicide ideation | 731 | Logistic | 0,89 | 0,76 , 1,05 | 0,17 |
| Psychotic symptoms | 1281 | Logistic | 1,15 | 1,03 , 1,29 | **1,40E-02** |
| Sleeping disturbances | 1228 | Logistic | 0,93 | 0,83 , 1,04 | 0,21 |

| | | | | | |
|---|---|---|---|---|---|
| **Sociodemographic and health phenotypes** | | | | | |
| *EuropASI* | | | | | |
| Legal status | 984 | Ordinal | 1,06 | 0,92 , 1,23 | 0,42 |
| Employment status | 984 | Ordinal | 1,10 | 0,99 , 1,23 | 0,08 |
| Medical status | 982 | Ordinal | 1,02 | 0,9 , 1,16 | 0,71 |
| Psychiatric status | 984 | Ordinal | 1,03 | 0,92 , 1,15 | 0,60 |
| Drug use | 984 | Ordinal | 0,96 | 0,86 , 1,08 | 0,48 |
| Alcohol use | 984 | Ordinal | 1,03 | 0,92 , 1,16 | 0,61 |
| Family/Social relationships | 981 | Ordinal | 1,01 | 0,91 , 1,13 | 0,82 |
| *36-Item Short Form Survey (SF-36)* | | | | | |
| Physical health | 751 | Linear | -0,53 | -1,26 , 0,21 | 0,16 |
| Mental health | 751 | Linear | -0,10 | -1,08 , 0,89 | 0,85 |
| Criminal record | 713 | Logistic | 1,07 | 0,92 , 1,24 | 0,41 |
| Unemployment | 1057 | Logistic | 1,13 | 1 , 1,28 | **4,71E-02** |
| Number of psychiatric hospitalizations | 760 | Negative binomial | 1,10 | 0,9 , 1,34 | 0,35 |
| Psychiatric family history | 840 | Logistic | 1,01 | 0,88 , 1,15 | 0,93 |
| Lifetime medical conditions | 1334 | Logistic | 0,99 | 0,88 , 1,11 | 0,86 |
| Substance use family history | 818 | Logistic | 1,03 | 0,9 , 1,19 | 0,64 |
| Educational attainment | 1333 | Ordinal | 1,01 | 1,11 , 0,91 | 0,87 |

[a] For binary traits sample size was calculated with the formula 4/(1/n1+1/n2)

[b] OR is reported for logistic regression and ordinal regression; Beta is reported for lineal regression; IRR is reported for negative binomial regression

[c] Logarithmic transformations were applied to continuous variables not following a normal distribution

**Supplementary Table 2g**. Association between the GPS for risk tolerance and 39 clinical variables from the SUD phenome. In bold nominal significant results.

| Phenotypes | *n* [a] | Regression | Estimate [b] | 95% CI | *p* |
|---|---|---|---|---|---|
| **SUD-related phenotypes** | | | | | |
| Age at onset of substance use [c] | 1308 | Linear | -0,01 | -0,02 , 3,00E-03 | 0,12 |
| Age at onset of SUD [c] | 1280 | Linear | -0,01 | -0,03 , 4,00E-03 | 0,14 |
| Years between substance use and SUD | 1280 | Linear | 0,99 | 0,9 , 1,08 | 0,77 |
| Years of substance use as proportion of lifespan | 1199 | Linear | 0,42 | -0,63 , 1,47 | 0,43 |
| Number of substances consumed | 879 | Negative binomial | 1,02 | 0,98 , 1,06 | 0,31 |
| Number of therapeutic community interventions | 1345 | Negative binomial | 1,02 | 0,93 , 1,12 | 0,69 |
| Number of inpatient detoxifications | 1327 | Negative binomial | 1,01 | 0,9 , 1,13 | 0,88 |
| Number of outpatient treatments | 1252 | Negative binomial | 1,06 | 1 , 1,12 | **4,33E-02** |
| **Comorbidity and personality traits** | | | | | |
| *Mental disorders in DSM-IV* | | | | | |
| Borderline personality disorder | 447 | Logistic | 0,97 | 0,8 , 1,18 | 0,76 |
| Major depressive disorder | 828 | Logistic | 0,89 | 0,77 , 1,02 | 0,09 |
| Antisocial personality disorder | 560 | Logistic | 0,99 | 0,83 , 1,17 | 0,87 |
| Psychotic disorder | 593 | Logistic | 1,08 | 0,84 , 1,38 | 0,55 |
| Anxiety disorder | 654 | Logistic | 1,03 | 0,89 , 1,21 | 0,69 |
| Attention deficit hyperactivity disorder | 700 | Logistic | 1,08 | 0,93 , 1,26 | 0,30 |
| *Zuckerman–Kuhlman Personality Questionnaire (ZKPQ)* | | | | | |
| Neuroticism Anxiety personality factor | 663 | Linear | -0,51 | -0,88 , -0,14 | **6,90E-03** |
| Aggression Hostility personality factor | 667 | Linear | 0,17 | -0,07 , 0,41 | 0,16 |
| Sociability personality factor | 632 | Linear | 0,09 | -0,18 , 0,36 | 0,51 |
| Impulsive sensation seeking personality factor | 666 | Linear | 0,32 | 0 , 0,65 | **4,98E-02** |
| Activity personality factor | 665 | Linear | 0,10 | -0,17 , 0,36 | 0,48 |
| Suicide attempt | 618 | Logistic | 1,11 | 0,95 , 1,29 | 0,18 |
| Suicide ideation | 731 | Logistic | 1,04 | 0,88 , 1,23 | 0,62 |
| Psychotic symptoms | 1281 | Logistic | 1,00 | 0,89 , 1,12 | 0,98 |
| Sleeping disturbances | 1228 | Logistic | 1,09 | 0,97 , 1,22 | 0,16 |

| | | | | | |
|---|---|---|---|---|---|
| **Sociodemographic and health phenotypes** | | | | | |
| *EuropASI* | | | | | |
| Legal status | 984 | Ordinal | 1,18 | 1,02 , 1,36 | **2,29E-02** |
| Employment status | 984 | Ordinal | 0,95 | 0,85 , 1,06 | 0,35 |
| Medical status | 982 | Ordinal | 1,03 | 0,91 , 1,16 | 0,64 |
| Psychiatric status | 984 | Ordinal | 0,97 | 0,87 , 1,08 | 0,59 |
| Drug use | 984 | Ordinal | 1,01 | 0,91 , 1,13 | 0,84 |
| Alcohol use | 984 | Ordinal | 1,03 | 0,92 , 1,15 | 0,58 |
| Family/Social relationships | 981 | Ordinal | 1,01 | 0,9 , 1,12 | 0,92 |
| *36-Item Short Form Survey (SF-36)* | | | | | |
| Physical health | 751 | Linear | 0,08 | -0,65 , 0,81 | 0,83 |
| Mental health | 751 | Linear | 0,30 | -0,67 , 1,27 | 0,55 |
| Criminal record | 713 | Logistic | 1,16 | 1 , 1,36 | 0,06 |
| Unemployment | 1057 | Logistic | 0,99 | 0,87 , 1,11 | 0,83 |
| Number of psychiatric hospitalizations | 760 | Negative binomial | 0,87 | 0,71 , 1,07 | 0,18 |
| Psychiatric family history | 840 | Logistic | 1,01 | 0,88 , 1,17 | 0,84 |
| Lifetime medical conditions | 1334 | Logistic | 1,05 | 0,94 , 1,18 | 0,37 |
| Substance use family history | 818 | Logistic | 1,11 | 0,96 , 1,28 | 0,15 |
| Educational attainment | 1333 | Ordinal | 1,04 | 1,15 , 0,94 | 0,42 |

[a] For binary traits sample size was calculated with the formula 4/(1/n1+1/n2)

[b] OR is reported for logistic regression and ordinal regression; Beta is reported for lineal regression;  IRR is reported for negative binomial regression

[c] Logarithmic transformations were applied to continuous variables not following a normal distribution

**Supplementary Table 2h**. Association between the GPS for suicide attempt and 39 clinical variables from the SUD phenome. In bold nominal significant results.

| Phenotypes | *n* [a] | Regression | Estimate [b] | 95% CI | *p* |
|---|---|---|---|---|---|
| **SUD-related phenotypes** | | | | | |
| Age at onset of substance use [c] | 1308 | Linear | -0,01 | -0,03 , 2,00E-03 | 0,11 |
| Age at onset of SUD [c] | 1280 | Linear | -0,02 | -0,04 , -0,01 | **1,12E-02** |
| Years between substance use and SUD | 1280 | Linear | 0,92 | 0,84 , 1,01 | 0,08 |
| Years of substance use as proportion of lifespan | 1199 | Linear | 0,03 | -1,04 , 1,09 | 0,96 |
| Number of substances consumed | 879 | Negative binomial | 1,00 | 0,96 , 1,05 | 0,85 |
| Number of therapeutic community interventions | 1345 | Negative binomial | 0,96 | 0,87 , 1,06 | 0,40 |
| Number of inpatient detoxifications | 1327 | Negative binomial | 1,02 | 0,9 , 1,14 | 0,79 |
| Number of outpatient treatments | 1252 | Negative binomial | 1,06 | 1 , 1,12 | **4,10E-02** |
| **Comorbidity and personality traits** | | | | | |
| *Mental disorders in DSM-IV* | | | | | |
| Borderline personality disorder | 447 | Logistic | 1,10 | 0,9 , 1,35 | 0,34 |
| Major depressive disorder | 828 | Logistic | 1,08 | 0,93 , 1,25 | 0,33 |
| Antisocial personality disorder | 560 | Logistic | 1,18 | 0,99 , 1,4 | 0,07 |
| Psychotic disorder | 593 | Logistic | 1,09 | 0,85 , 1,42 | 0,50 |
| Anxiety disorder | 654 | Logistic | 1,03 | 0,88 , 1,21 | 0,69 |
| Attention deficit hyperactivity disorder | 700 | Logistic | 1,09 | 0,93 , 1,27 | 0,30 |
| *Zuckerman–Kuhlman Personality Questionnaire (ZKPQ)* | | | | | |
| Neuroticism Anxiety personality factor | 663 | Linear | 0,09 | -0,29 , 0,48 | 0,63 |
| Aggression Hostility personality factor | 667 | Linear | 0,26 | 0,02 , 0,51 | **3,60E-02** |
| Sociability personality factor | 632 | Linear | 0,04 | -0,24 , 0,31 | 0,80 |
| Impulsive sensation seeking personality factor | 666 | Linear | 0,00 | -0,33 , 0,34 | 0,99 |
| Activity personality factor | 665 | Linear | 0,24 | -0,03 , 0,52 | 0,09 |
| Suicide attempt | 618 | Logistic | 1,01 | 0,87 , 1,18 | 0,85 |
| Suicide ideation | 731 | Logistic | 1,16 | 0,98 , 1,37 | 0,08 |
| Psychotic symptoms | 1281 | Logistic | 1,15 | 1,03 , 1,29 | **1,69E-02** |
| Sleeping disturbances | 1228 | Logistic | 0,97 | 0,87 , 1,09 | 0,65 |

| | | | | | |
|---|---|---|---|---|---|
| **Sociodemographic and health phenotypes** | | | | | |
| *EuropASI* | | | | | |
| Legal status | 984 | Ordinal | 1,18 | 1,01 , 1,37 | **3,27E-02** |
| Employment status | 984 | Ordinal | 1,03 | 0,92 , 1,16 | 0,56 |
| Medical status | 982 | Ordinal | 1,14 | 1 , 1,29 | **4,93E-02** |
| Psychiatric status | 984 | Ordinal | 1,13 | 1,01 , 1,27 | **3,88E-02** |
| Drug use | 984 | Ordinal | 1,05 | 0,93 , 1,18 | 0,44 |
| Alcohol use | 984 | Ordinal | 1,02 | 0,91 , 1,15 | 0,75 |
| Family/Social relationships | 981 | Ordinal | 1,13 | 1 , 1,26 | 0,05 |
| *36-Item Short Form Survey (SF-36)* | | | | | |
| Physical health | 751 | Linear | -0,47 | -1,21 , 0,27 | 0,22 |
| Mental health | 751 | Linear | -0,19 | -1,18 , 0,8 | 0,71 |
| Criminal record | 713 | Logistic | 1,14 | 0,98 , 1,32 | 0,11 |
| Unemployment | 1057 | Logistic | 1,05 | 0,93 , 1,19 | 0,40 |
| Number of psychiatric hospitalizations | 760 | Negative binomial | 1,08 | 0,88 , 1,32 | 0,47 |
| Psychiatric family history | 840 | Logistic | 0,94 | 0,82 , 1,08 | 0,40 |
| Lifetime medical conditions | 1334 | Logistic | 1,13 | 1,01 , 1,27 | **3,84E-02** |
| Substance use family history | 818 | Logistic | 1,11 | 0,96 , 1,28 | 0,15 |
| Educational attainment | 1333 | Ordinal | 0,90 | 1 , 0,81 | **4,30E-02** |

[a] For binary traits sample size was calculated with the formula 4/(1/n1+1/n2)

[b] OR is reported for logistic regression and ordinal regression; Beta is reported for lineal regression;  IRR is reported for negative binomial regression

[c] Logarithmic transformations were applied to continuous variables not following a normal distribution

**Supplementary Table 2i.** Association between the GPS for educational attainment and 39 clinical variables from the SUD phenome. In bold nominal significant results.

| Phenotypes | *n* [a] | Regression | Estimate [b] | 95% CI | *p* |
|---|---|---|---|---|---|
| **SUD-related phenotypes** | | | | | |
| Age at onset of substance use [c] | 1308 | Linear | 0,02 | 0,01 , 0,03 | **8,11E-03** |
| Age at onset of SUD [c] | 1280 | Linear | 0,02 | 4,00E-03 , 0,04 | **1,48E-02** |
| Years between substance use and SUD | 1280 | Linear | 1,07 | 0,98 , 1,17 | 0,15 |
| Years of substance use as proportion of lifespan | 1199 | Linear | -0,73 | -1,77 , 0,31 | 0,17 |
| Number of substances consumed | 879 | Negative binomial | 1,00 | 0,96 , 1,04 | 0,89 |
| Number of therapeutic community interventions | 1345 | Negative binomial | 0,89 | 0,81 , 0,98 | **2,28E-02** |
| Number of inpatient detoxifications | 1327 | Negative binomial | 0,89 | 0,8 , 1 | 0,06 |
| Number of outpatient treatments | 1252 | Negative binomial | 0,91 | 0,87 , 0,97 | **1,64E-03** |
| **Comorbidity and personality traits** | | | | | |
| *Mental disorders in DSM-IV* | | | | | |
| Borderline personality disorder | 447 | Logistic | 0,96 | 0,79 , 1,18 | 0,70 |
| Major depressive disorder | 828 | Logistic | 0,91 | 0,79 , 1,06 | 0,24 |
| Antisocial personality disorder | 560 | Logistic | 0,90 | 0,75 , 1,08 | 0,24 |
| Psychotic disorder | 593 | Logistic | 1,04 | 0,8 , 1,37 | 0,76 |
| Anxiety disorder | 654 | Logistic | 0,97 | 0,83 , 1,14 | 0,71 |
| Attention deficit hyperactivity disorder | 700 | Logistic | 0,89 | 0,76 , 1,05 | 0,16 |
| *Zuckerman–Kuhlman Personality Questionnaire (ZKPQ)* | | | | | |
| Neuroticism Anxiety personality factor | 663 | Linear | -0,39 | -0,78 , -0,01 | **4,50E-02** |
| Aggression Hostility personality factor | 667 | Linear | -0,18 | -0,42 , 0,07 | 0,16 |
| Sociability personality factor | 632 | Linear | 0,10 | -0,18 , 0,38 | 0,47 |
| Impulsive sensation seeking personality factor | 666 | Linear | -0,05 | -0,39 , 0,28 | 0,75 |
| Activity personality factor | 665 | Linear | 0,17 | -0,11 , 0,45 | 0,24 |
| Suicide attempt | 618 | Logistic | 1,03 | 0,89 , 1,2 | 0,66 |
| Suicide ideation | 731 | Logistic | 1,00 | 0,85 , 1,17 | 0,96 |
| Psychotic symptoms | 1281 | Logistic | 0,91 | 0,82 , 1,02 | 0,12 |
| Sleeping disturbances | 1228 | Logistic | 0,91 | 0,81 , 1,02 | 0,09 |

| | | | | | |
|---|---|---|---|---|---|
| **Sociodemographic and health phenotypes** | | | | | |
| *EuropASI* | | | | | |
| Legal status | 984 | Ordinal | 0,96 | 0,82 , 1,11 | 0,55 |
| Employment status | 984 | Ordinal | 0,90 | 0,81 , 1,01 | 0,07 |
| Medical status | 982 | Ordinal | 1,00 | 0,88 , 1,13 | 0,99 |
| Psychiatric status | 984 | Ordinal | 0,91 | 0,81 , 1,01 | 0,08 |
| Drug use | 984 | Ordinal | 0,95 | 0,84 , 1,06 | 0,36 |
| Alcohol use | 984 | Ordinal | 0,98 | 0,87 , 1,1 | 0,75 |
| Family/Social relationships | 981 | Ordinal | 0,86 | 0,77 , 0,96 | **1,00E-02** |
| *36-Item Short Form Survey (SF-36)* | | | | | |
| Physical health | 751 | Linear | 0,53 | -0,24 , 1,3 | 0,18 |
| Mental health | 751 | Linear | 0,59 | -0,43 , 1,62 | 0,26 |
| Criminal record | 713 | Logistic | 0,67 | 0,57 , 0,78 | **8,03E-07** |
| Unemployment | 1057 | Logistic | 0,82 | 0,72 , 0,92 | **1,34E-03** |
| Number of psychiatric hospitalizations | 760 | Negative binomial | 1,15 | 0,94 , 1,41 | 0,17 |
| Psychiatric family history | 840 | Logistic | 1,08 | 0,94 , 1,23 | 0,28 |
| Lifetime medical conditions | 1334 | Logistic | 0,99 | 0,88 , 1,11 | 0,88 |
| Substance use family history | 818 | Logistic | 0,80 | 0,7 , 0,92 | **2,20E-03** |
| Educational attainment | 1333 | Ordinal | 1,39 | 1,54 , 1,25 | **3,34E-10** |

[a] For binary traits sample size was calculated with the formula 4/(1/n1+1/n2)

[b] OR is reported for logistic regression and ordinal regression; Beta is reported for lineal regression; IRR is reported for negative binomial regression

[c] Logarithmic transformations were applied to continuous variables not following a normal distribution

**Supplementary Table 2j**. Association between the GPS for well-being and 39 clinical variables from the SUD phenome. In bold nominal significant results.

| Phenotypes | *n* [a] | Regression | Estimate [b] | 95% CI | *p* |
|---|---|---|---|---|---|
| **SUD-related phenotypes** | | | | | |
| Age at onset of substance use [c] | 1308 | Linear | -0,01 | -0,02 , 3,00E-03 | 0,13 |
| Age at onset of SUD [c] | 1280 | Linear | -0,01 | -0,03 , 0,01 | 0,25 |
| Years between substance use and SUD | 1280 | Linear | 0,98 | 0,9 , 1,07 | 0,69 |
| Years of substance use as proportion of lifespan | 1199 | Linear | 0,65 | -0,4 , 1,7 | 0,23 |
| Number of substances consumed | 879 | Negative binomial | 0,99 | 0,95 , 1,04 | 0,81 |
| Number of therapeutic community interventions | 1345 | Negative binomial | 0,99 | 0,9 , 1,08 | 0,77 |
| Number of inpatient detoxifications | 1327 | Negative binomial | 0,99 | 0,88 , 1,11 | 0,87 |
| Number of outpatient treatments | 1252 | Negative binomial | 0,91 | 0,86 , 0,96 | **7,00E-04** |
| **Comorbidity and personality traits** | | | | | |
| *Mental disorders in DSM-IV* | | | | | |
| Borderline personality disorder | 447 | Logistic | 1,07 | 0,87 , 1,31 | 0,52 |
| Major depressive disorder | 828 | Logistic | 0,99 | 0,86 , 1,15 | 0,90 |
| Antisocial personality disorder | 560 | Logistic | 1,06 | 0,9 , 1,26 | 0,49 |
| Psychotic disorder | 593 | Logistic | 0,85 | 0,65 , 1,11 | 0,23 |
| Anxiety disorder | 654 | Logistic | 0,90 | 0,76 , 1,06 | 0,20 |
| Attention deficit hyperactivity disorder | 700 | Logistic | 0,92 | 0,79 , 1,07 | 0,28 |
| *Zuckerman–Kuhlman Personality Questionnaire (ZKPQ)* | | | | | |
| Neuroticism Anxiety personality factor | 663 | Linear | -0,34 | -0,72 , 0,04 | 0,08 |
| Aggression Hostility personality factor | 667 | Linear | 0,05 | -0,2 , 0,29 | 0,71 |
| Sociability personality factor | 632 | Linear | 0,27 | -0,01 , 0,54 | 0,06 |
| Impulsive sensation seeking personality factor | 666 | Linear | 0,25 | -0,08 , 0,58 | 0,14 |
| Activity personality factor | 665 | Linear | 0,11 | -0,16 , 0,38 | 0,43 |
| Suicide attempt | 618 | Logistic | 0,95 | 0,82 , 1,1 | 0,50 |
| Suicide ideation | 731 | Logistic | 0,93 | 0,79 , 1,09 | 0,36 |
| Psychotic symptoms | 1281 | Logistic | 0,93 | 0,83 , 1,04 | 0,20 |
| Sleeping disturbances | 1228 | Logistic | 0,91 | 0,82 , 1,02 | 0,12 |

| | | | | | |
|---|---|---|---|---|---|
| **Sociodemographic and health phenotypes** | | | | | |
| *EuropASI* | | | | | |
| Legal status | 984 | Ordinal | 0,95 | 0,82 , 1,1 | 0,51 |
| Employment status | 984 | Ordinal | 0,95 | 0,85 , 1,06 | 0,37 |
| Medical status | 982 | Ordinal | 0,95 | 0,83 , 1,07 | 0,40 |
| Psychiatric status | 984 | Ordinal | 0,87 | 0,78 , 0,97 | **1,43E-02** |
| Drug use | 984 | Ordinal | 0,98 | 0,87 , 1,09 | 0,67 |
| Alcohol use | 984 | Ordinal | 1,01 | 0,9 , 1,14 | 0,83 |
| Family/Social relationships | 981 | Ordinal | 0,83 | 0,74 , 0,93 | **1,00E-03** |
| *36-Item Short Form Survey (SF-36)* | | | | | |
| Physical health | 751 | Linear | 0,64 | -0,1 , 1,37 | 0,09 |
| Mental health | 751 | Linear | 0,51 | -0,47 , 1,49 | 0,31 |
| Criminal record | 713 | Logistic | 0,90 | 0,78 , 1,05 | 0,17 |
| Unemployment | 1057 | Logistic | 0,88 | 0,77 , 0,99 | **3,59E-02** |
| Number of psychiatric hospitalizations | 760 | Negative binomial | 0,78 | 0,64 , 0,95 | **1,33E-02** |
| Psychiatric family history | 840 | Logistic | 0,90 | 0,79 , 1,03 | 0,13 |
| Lifetime medical conditions | 1334 | Logistic | 0,98 | 0,87 , 1,1 | 0,70 |
| Substance use family history | 818 | Logistic | 0,83 | 0,73 , 0,96 | **1,13E-02** |
| Educational attainment | 1333 | Ordinal | 1,06 | 1,18 , 0,96 | 0,23 |

[a] For binary traits sample size was calculated with the formula 4/(1/n1+1/n2)

[b] OR is reported for logistic regression and ordinal regression; Beta is reported for lineal regression;  IRR is reported for negative binomial regression

[c] Logarithmic transformations were applied to continuous variables not following a normal distribution

**Supplementary Table 3**.

*Interaction between GPSs and lifetime emotional. physical and/or sexual abuse in the SUD phenome*

| GPS | SUD-Phenome | *n* [a] | Estimate | 95% CI | *p* |
|---|---|---|---|---|---|
| Attention-deficit hyperactivity disorder | Age at onset of substance use [b] | 701 | -.002 | -.04 , .04 | .94 |
| | Years of substance use as proportion of lifespan | 631 | .32 | -2.57 , 3.2 | .83 |
| | Antisocial personality disorder | 358 | 1.00 | .66 , 1.51 | .99 |
| | Attention deficit hyperactivity disorder | 434 | .94 | .63 , 1.4 | .77 |
| | Educational attainment | 701 | .96 | .73 , 1.26 | .77 |
| Anxiety | Number of outpatient treatments | 681 | 1.06 | .92 , 1.22 | .42 |
| | Psychotic disorder | 180 | 1.10 | .61 , 1.99 | .76 |
| | SF36 Physical health | 509 | .43 | -1.27 , 2.12 | .62 |
| | Psychiatric family history | 524 | 1.03 | .73 , 1.46 | .86 |
| Bipolar disorder | Psychotic symptoms | 678 | 1.01 | .74 , 1.39 | .93 |
| | Unemployment | 568 | 1.08 | .78 , 1.51 | .64 |
| | Number of psychiatric hospitalizations | 504 | .87 | .53 , 1.42 | .57 |
| | Substance use family history | 516 | .98 | .69 , 1.39 | .91 |
| Depression | Years between substance use and SUD | 691 | .96 | .74 , 1.25 | .78 |
| | Number of outpatient treatments | 681 | 1.07 | .92 , 1.23 | .39 |
| | ZKPQ-Neuroticism Anxiety personality factor | 492 | .20 | -.64 , 1.05 | .64 |
| | ZKPQ-Aggression Hostility personality factor | 495 | -.28 | -.83 , .26 | .31 |

| | | | | | |
|---|---|---|---|---|---|
| | Suicide attempt | 725 | .93 | .69 , 1.26 | .65 |
|  | Criminal record | 488 | .76 | .53 , 1.1 | .14 |
| | Psychiatric family history | 524 | 1.31 | .92 , 1.86 | .13 |
| Post-traumatic stress disorder | Age at onset of substance use [b] | 701 | -.02 | -.06 , .03 | .45 |
| | Number of inpatient detoxifications | 702 | .91 | .65 , 1.26 | .56 |
| | Unemployment | 568 | .85 | .6 , 1.21 | .38 |
| | Educational attainment | 701 | 1.08 | .81 , 1.44 | .58 |
| Schizophrenia | Age at onset of substance use [b] | 701 | .03 | -.01 , .08 | .11 |
| | Psychotic disorder | 180 | .85 | .45 , 1.59 | .60 |
| | Psychotic symptoms | 678 | .97 | .7 , 1.33 | .83 |
| | Unemployment | 568 | .76 | .54 , 1.07 | .12 |
| Risk tolerance | Number of outpatient treatments | 681 | 1.12 | .96 , 1.31 | .14 |
| | ZKPQ-Neuroticism Anxiety personality factor | 492 | .29 | -.6 , 1.18 | .53 |
|  | ZKPQ-Impulsive sensation seeking personality factor | 493 | .39 | -.37 , 1.14 | .32 |
| | EuropASI-Legal status | 701 | 1.07 | .75 , 1.52 | .72 |
| | Unemployment | 568 | .85 | .6 , 1.21 | .38 |
| Suicide attempt | Age at onset of SUD [b] | 699 | .00 | -.04 , .05 | .89 |
| | Number of outpatient treatments | 681 | .98 | .84 , 1.13 | .77 |
|  | ZKPQ-Aggression Hostility personality factor | 495 | -.02 | -.58 , .55 | .96 |
| | Psychotic symptoms | 678 | 1.03 | .75 , 1.41 | .86 |
| | EuropASI-Legal status | 701 | 1.08 | .76 , 1.55 | .66 |

| | | | | | |
|---|---|---|---|---|---|
| | EuropASI-Medical status | 699 | 1.13 | .84 , 1.51 | .41 |
| | EuropASI-Psychiatric status | 701 | 1.35 | 1.03 , 1.78 | **2.94E-02** |
| | Lifetime medical conditions | 705 | .95 | .7 , 1.31 | .76 |
| | Educational attainment | 701 | 1.10 | .83 , 1.46 | .51 |
| Educational attainment | Age at onset of substance use [b] | 701 | .01 | -.03 , .06 | .50 |
| | Age at onset of SUD [b] | 699 | .01 | -.03 , .06 | .54 |
| | Number of therapeutic community interventions | 698 | .86 | .67 , 1.11 | .26 |
| | Number of outpatient treatments | 681 | 1.06 | .91 , 1.23 | .47 |
| | ZKPQ-Neuroticism Anxiety personality factor | 492 | .16 | -.73 , 1.06 | .72 |
| | Criminal record | 488 | 1.27 | .86 , 1.89 | .23 |
| | Unemployment | 568 | .73 | .51 , 1.04 | .08 |
| | Substance use family history | 516 | 1.00 | .7 , 1.45 | .99 |
| | Educational attainment | 701 | .94 | .71 , 1.26 | .69 |
| Well being | Number of outpatient treatments | 681 | 1.00 | .86 , 1.17 | .97 |
| | EuropASI- Psychiatric status | 701 | 1.01 | .77 , 1.32 | .96 |
| | EuropASI-Family/social relationships | 701 | .95 | .73 , 1.25 | .22 |
| | Unemployment | 568 | .87 | .61 , 1.25 | .46 |
| | Number of psychiatric hospitalizations | 504 | 1.23 | .73 , 2.07 | .44 |
| | Substance use family history | 516 | 1.05 | .73 , 1.52 | .80 |

*Note.* Odds Ratio (OR) is reported for logistic regression and ordinal regression; Beta is reported for lineal regression; Incidence Rate Ratio (IRR) is reported for negative binomial regression.

[a] For binary traits sample size was calculated with the formula 4/(1/n1+1/n2).

[b] Logarithmic transformations were applied to continuous variables not following a normal distribution.
